# Lifetime incidence and age of onset of mental disorders, and 12-month service utilization in primary and secondary care: a Finnish nationwide registry study

**DOI:** 10.1101/2024.12.04.24318482

**Authors:** Kimmo Suokas, Ripsa Niemi, Mai Gutvilig, John J. McGrath, Kaisla Komulainen, Jaana Suvisaari, Marko Elovainio, Sonja Lumme, Sami Pirkola, Christian Hakulinen

## Abstract

Previous studies have estimated lifetime incidence, age-specific incidence, age of onset, and service utilization for mental disorders but none have used nationwide data from both primary and secondary care. This study used nationwide Finnish data (2000–2020), including both care settings for the first time. We followed 6.4 million individuals for 98.5 million person-years, calculating cumulative incidence while accounting for competing risks. By age 100, lifetime incidence of any diagnosed mental disorder was 76.7% (95% CI, 76.6–76.7) in women and 69.7% (69.6–69.8) in men. At age 75, stricter estimates for non-organic disorders (ICD-10: F10–F99) were 65.6% (65.5–65.7) for women and 60.0% (59.9–60.1). Anxiety disorders (F40–F48) had the highest cumulative incidence. Median age of onset of non–organic mental disorders was 24.1 (interquartile range 14.8–43.3) in women and 20.0 (7.3–42.2) in men. Service utilization within 12 months was 9.0% for women and 7.7% for men. Most, though not all, individuals experience at least one type of mental disorder, often during youth. Capturing the overall occurrence of mental disorders requires including both primary and secondary care data.

## Introduction

Mental disorders are prevalent, commonly have their first onset in childhood and adolescence, tend to shift from one to another, and thus constitute a major source of years lived with disability throughout the life course [1–5]. Understanding the fundamental aspects of lifetime incidence, age of onset, and service utilization for different mental disorders may help conceptualize mental disorders, identify windows for interventions, and plan efficient services.

Recent findings indicate that mental disorders eventually affect almost everyone [3]. In a Danish register-study combining data on secondary care and psychotropic medication prescriptions (as a proxy marker for a diagnosis of mental disorders), the lifetime cumulative incidence of a mental disorder was 82.6% by the age of 100 [1]. On the other hand, major survey studies have estimated that approximately half of the population experience a mental disorder by the age of 75 years [2, 6]. Previous studies on lifetime incidence differ regarding the age considered—75, 80, or 100 years—and whether organic and other neuropsychiatric diagnoses were included, contributing to variations in reported lifetime estimates [2, 7, 8]. In Finland, where the median age at death is currently 85 for women and 78 for men, it has been estimated that over 20% of women born after 1975 will live beyond 100 years, indicating the relevance of cumulative incidence estimates at various ages [9, 10]. To date, there are no nationwide estimates of lifetime incidence of all diagnosed mental disorders in both primary and secondary care.

Similarly, several studies have evaluated the age of onset of mental disorders, but comprehensive nationwide reports are lacking so far. Based on a meta-analysis of survey studies, incidence peaks at the age of 14.5 years [11]. There is substantial variation in the peak age of onset by gender and diagnosis, with the traditional childhood-onset disorders showing the earliest age of onset and organic mental disorders the latest [2, 7, 8, 11, 12].

Mental disorders cause remarkable burden throughout life with varying patterns of remission and relapse [13, 14], and estimates for the prevalence of mental disorders vary between studies [15, 16]. Age- and diagnosis-specific analysis of medical service utilization captures both incident and chronic or recurrent cases, and together with incidence data may provide information on the overall need of care for different disorders throughout the life-course.

The aims of the present study were to estimate lifetime and age-specific cumulative incidence, the age of onset, and 12-month age-specific overall and diagnosis-specific service utilization for diagnosed mental disorders using nationwide population-based register data, covering both primary and secondary care.

## Methods

This register-based cohort study included all individuals born in Finland or elsewhere, from January 1, 1900, through December 31, 2019, and present in the Finnish population register at some point between January 1, 2000, and December 31, 2020.

The Research Ethics Committee of the Finnish Institute for Health and Welfare approved the study protocol (decision #10/2016§751). Data were linked with permission from Statistics Finland (TK–53–1696-16) and the Finnish Institute of Health and Welfare. Informed consent is not required for register-based studies in Finland.

### Data Sources

Data on the time of birth, death, and permanent emigration from Finland were extracted from the population register of Statistics Finland, which includes data on the total population on the last day of each study year.

Information on healthcare contacts was obtained from the Finnish Care Register for Health Care and the Register of Primary Health Care Visits, which show good consistence and adequate diagnostic reliability [17]. Psychiatric inpatient care can be dependably recognized since 1975, secondary outpatient care has been included since 1998 and primary care since 2011 [18].

The *International Statistical Classification of Diseases and Related Health Problems, Tenth Revision* (*ICD–10*) has been used in Finland since 1996. Prior to that, the Finnish version of the ICD–9 was used from 1987 to 1995, and ICD–8 from 1969 to 1986. In some primary care facilities, the International Classification of Primary Care, Second Edition (ICPC–2), is used instead of ICD–10. These diagnoses were converted to corresponding ICD–10 sub-chapter categories, and the registers were pre-processed for maximizing the accuracy of the data [18].

### Study Design

The primary estimate of an incident mental disorder included a diagnosis of any mental health disorder at inpatient or outpatient secondary services, or in primary care. In addition, we examined diagnosis-specific incidence and service utilization for ICD–10 sub-chapter categories and a wide range of particular diagnostic categories.

Persons were followed from January 1, 2000, or the earliest age at which a person might possibly develop the specific disorder (35 years for organic mental disorders, 1 year for disorders with onset commonly in childhood, and 5 years for others, Supplementary Table 1), whichever came later. The follow-up ended at the first recorded medical contact with the mental disorder diagnosis, 100th birthday, death, permanent emigration from Finland, or December 31, 2020, whichever came first. We excluded disorder-specific prevalent cases at the start of the follow-up period, which included those with inpatient treatments between 1975 and 1999, and those with outpatient care between 1998 and 1999.

Individuals aged under 100 with at least one medical contact for a diagnosed mental disorder in 2019 were identified to calculate the 12-month service utilization rate. This includes both first-time and prevalent cases, showing the number of people with medical contacts due to mental health conditions that year. The denominator included all individuals under 100 in the Finnish population register as of December 31, 2019. The year 2019 was chosen as the most recent year with data available before the COVID-19 pandemic.

### Statistical Analysis

Cumulative incidence estimates the percentage of individuals diagnosed with a mental disorder by a certain age using the Aalen–Johansen estimator, from the earliest possible onset age to the 100th birthday. The cumulative incidence at age 100 estimate the lifetime risk. Death or emigration from Finland were considered as competing risks. The cumulative incidence estimates were calculated for any disorder in the whole population, separately for men and women and separately for specific mental disorders.

Incidence rates across the age range depict the number of people with a first-time mental disorder diagnosis per 10 000 person-years at risk. The incidence rates were estimated at 1-year age intervals to evaluate the most common age to receive a first-time diagnosis. The 95% confidence intervals (CI) were estimated using Poisson regression. Median age of onset was defined as the age at which half of the lifetime incidence was reached [7].

Service utilization depicts the percentage of individuals aged under 100 years who had a health care visit with a mental disorder diagnosis in the year 2019 and were included in the study population on December 31, 2019. This was estimated in 1-year age intervals and for all ages.

We conducted two sensitivity analyses with stricter criteria for identifying prevalent cases to assess the robustness of our lifetime incidence estimates. First, we shortened the study period to 2003–2020, introducing an additional three-year washout period for inpatient and secondary outpatient treatments and a one-year retrospective washout for primary care; individuals with any primary care contacts on the year 2011 were excluded at the beginning of follow-up. This excluded individuals with recurrent primary care contacts but introduced potential immortal time and selection bias, as only those at risk of a first primary care contact until 2011 were eligible for exclusion. Second, we restricted follow-up to 2012–2020, with washout periods from 1975–2011 for inpatient care, 1998–2011 for secondary outpatient care, and 2011 for primary care. This avoided immortal time and selection bias but shortened the study duration. Further details are available in Supplementary Fig. 1.

Analyses were conducted using R version 4.2.2 (The R Foundation).

## Results

Altogether, 6 356 053 Finnish residents were followed for 98.5 million person-years. A total of 1 737 004 persons had their first healthcare contact for any mental disorder during the follow-up, 600 319 persons died, and 157 811 persons were censored due to emigration. Numbers of individuals within each diagnostic category are presented in Supplementary Table 1. Fig. 1 shows the overall cumulative incidence, incidence rate, and 12-month service utilization for all mental disorders; Figs. 2–4 show the corresponding estimates by ICD–10 sub-chapter category. Results for all specific diagnoses can be seen in interactive online material at https://mentalnet.shinyapps.io/lifetime/.

**Fig. 1:**
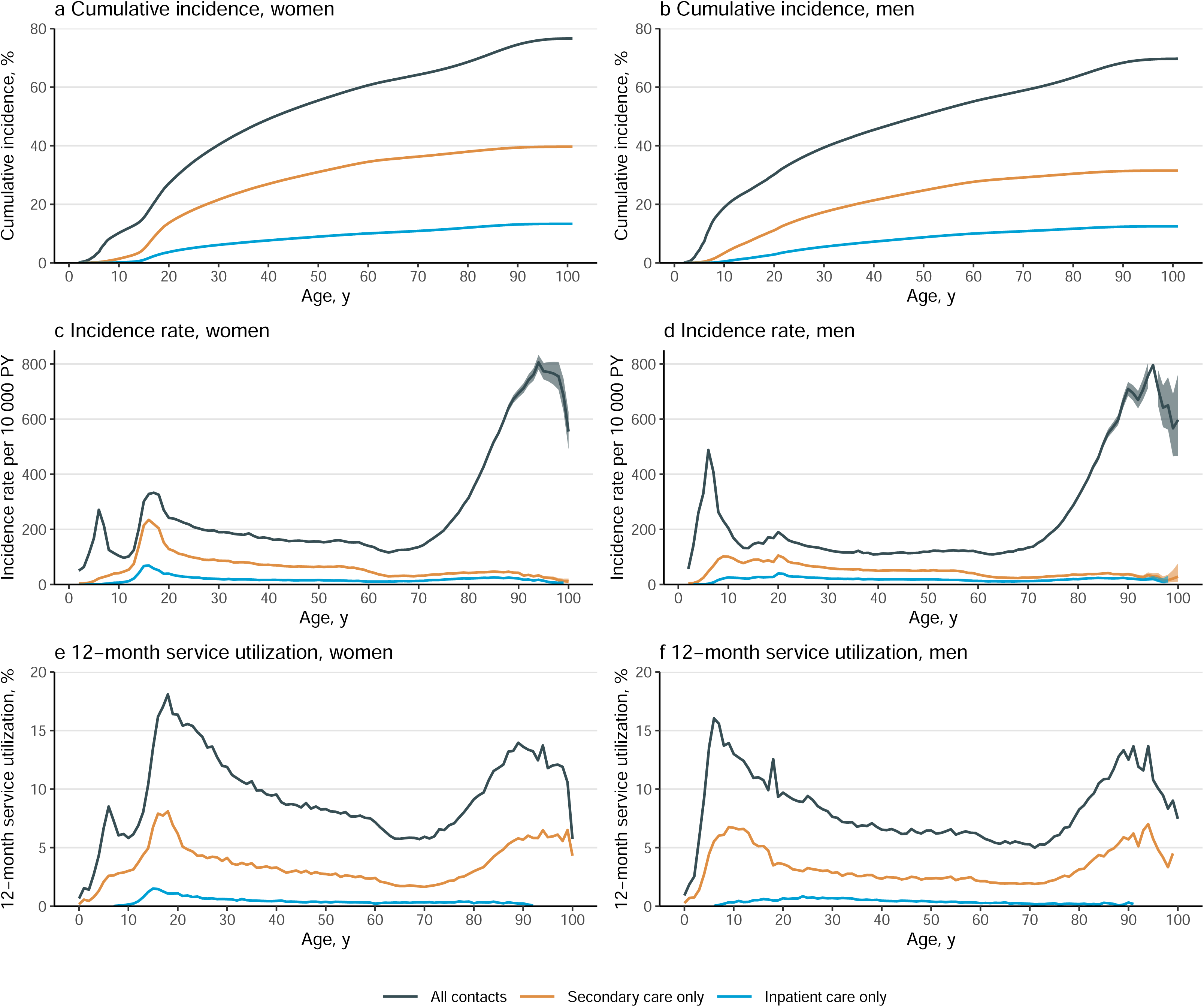
Cumulative incidence, incidence rate, and 12-month service utilization of mental disorders by gender and treatment type ^1^ Service utilization is the number of individuals with any medical contacts with a diagnosis of a mental disorders during the year 2019, divided by the number of individuals in the study population on December 31, 2019.

**Fig. 2:**
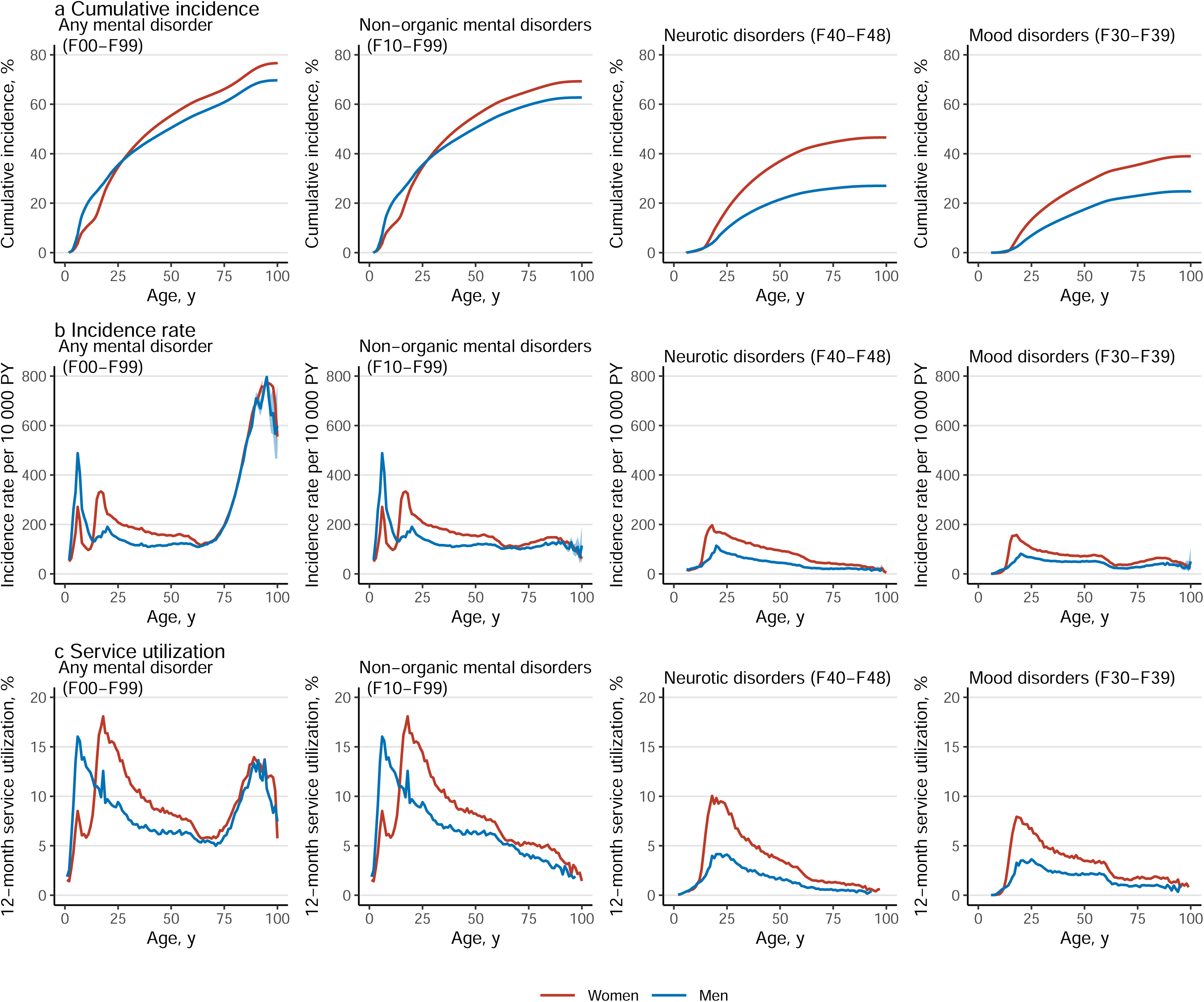
Cumulative incidence, incidence rate, and 12-month service utilization of mental disorders by gender and diagnosis ^1^ Service utilization is the number of individuals with any medical contacts with a diagnosis of a mental disorders during the year 2019, divided by the number of individuals in the study population on December 31, 2019.

### Cumulative Incidence

Cumulative incidence of any diagnosed mental disorder (ICD–10: F00–F99) at the age 100 years was 76.7% (76.6–76.7) for women and 69.7% (69.6–69.8) for men (Fig. 1a and 1b, and Table 1); in secondary care, it was 39.7% (39.6–39.8) for women and 31.5% (31.4–31.6) for men (Fig. 1a and 1b, and Supplementary Table 2); and in psychiatric inpatient care, 13.3% (13.3–13.4) for women and 12.5% (12.4–12.5) for men (Fig. 1a and 1b, and Supplementary Table 3). When organic mental disorders were excluded, cumulative incidence of any diagnosed mental disorder (F10–F99) at the age 100 and 75 years reduced to 69.3% (69.2–69.4) and 65.6% (65.5–65.7) in women and to 62.7% (62.6–62.8) and 60.0% (59.9–60.1) in men, respectively (Fig. 2a). Neurotic, stress–related and somatoform disorders (F40–F48) showed the highest cumulative incidence in women (46.6% [46.5–46.7]) and in men (27.0% [26.9–27.1]); Table 1 shows the cumulative incidence estimates for each ICD–10 sub-chapter category at different ages. Corresponding estimates for psychiatric secondary care and psychiatric inpatient care alone and a range of more detailed diagnostic categories are shown in Supplementary Tables 2–4.

**Table 1:**
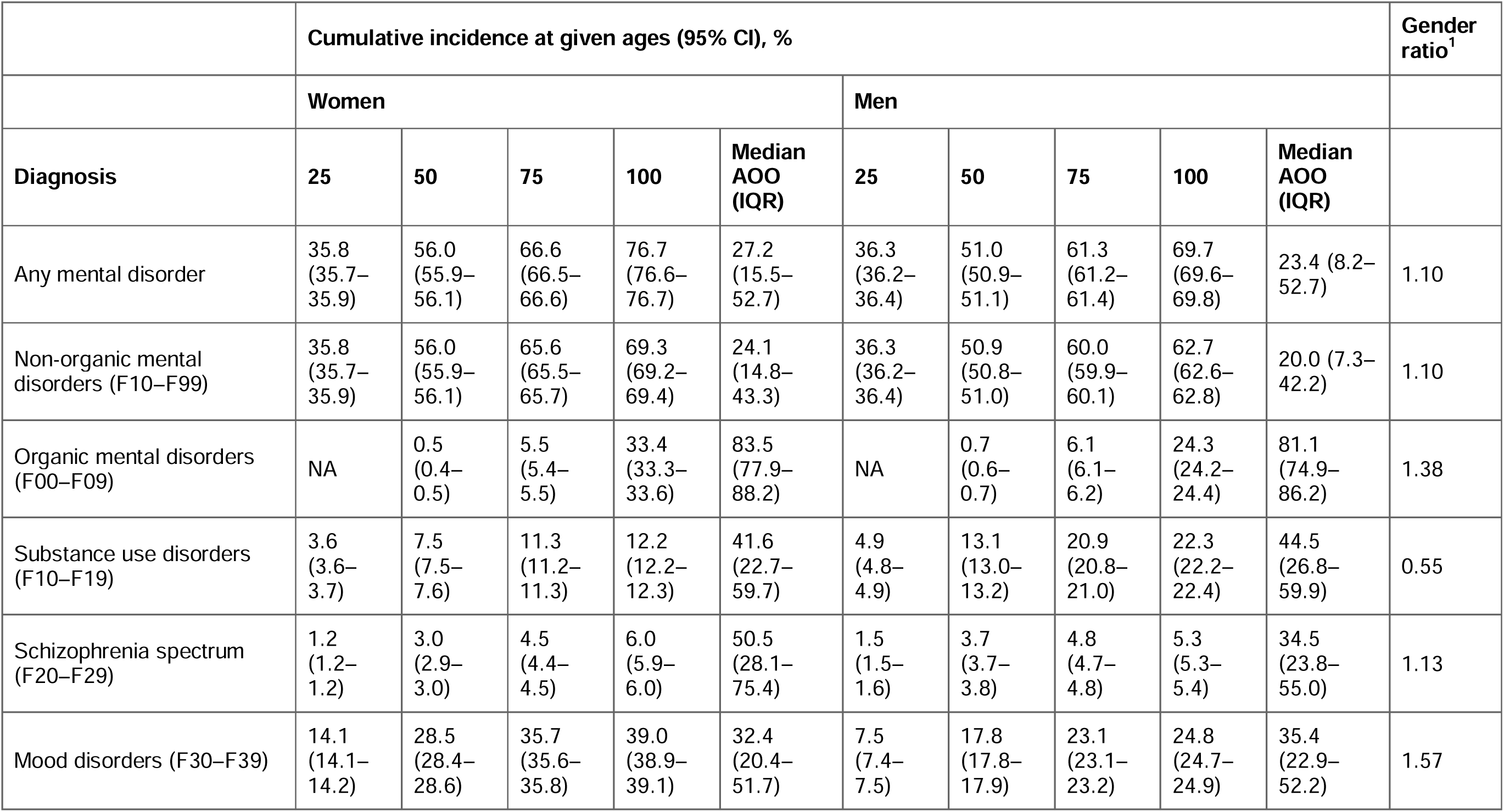
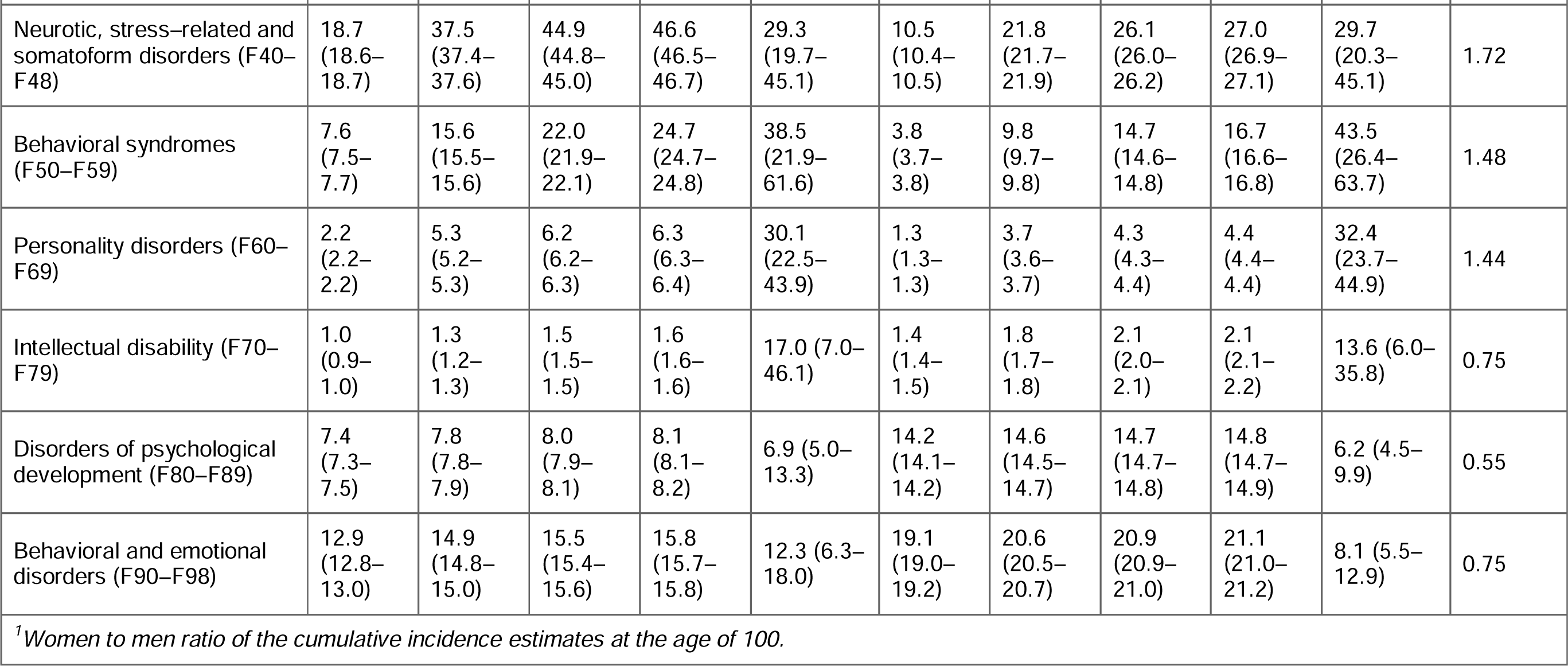
Cumulative incidence of mental disorders at the ages of 25, 50, 75, and 100 years, and median age of onset (AOO) and interquartile range (IQR) by gender and ICD–10 sub-chapter category

Cumulative incidence of any mental disorder was higher in men than in women until the age of 26.2 years, when the value was 37.3% (37.2–37.4) for women and men (Fig. 2a). Thereafter, cumulative incidence was higher in women. Behavioral and emotional disorders with onset usually occurring in childhood and adolescence (F90–F98) and disorders of psychological development (F80–F89) were the most common ICD–10 sub-chapter categories in early life; neurotic, stress–related and somatoform disorders (F40–F48) became the sub-chapter category with the highest cumulative incidence at the age of 21 in women and 46 in men and remained thereafter (Supplementary Fig. 2).

### Incidence Rates and Age of Onset Curves

The age-specific incidence rates in childhood and adolescence showed a bimodal pattern (Fig. 1c and 1d). The first peak in incidence was at the age of 6 in both boys and girls with most diagnoses from the sub-chapter categories of disorders of psychological development (F80–F89) and behavioral and emotional disorders (F90–F98) (Figs. 3b and 4b). The second peak was at the age of 15–18 in girls and outweighed the first peak, whereas in boys, the second peak was at the age of 20 and was much smaller than the first one. The two most prominent diagnoses at the second peak were mood disorders (F30–F39) and neurotic, stress–related and somatoform disorders (F40–F48) in both girls and boys (Fig. 2b).

**Fig. 3:**
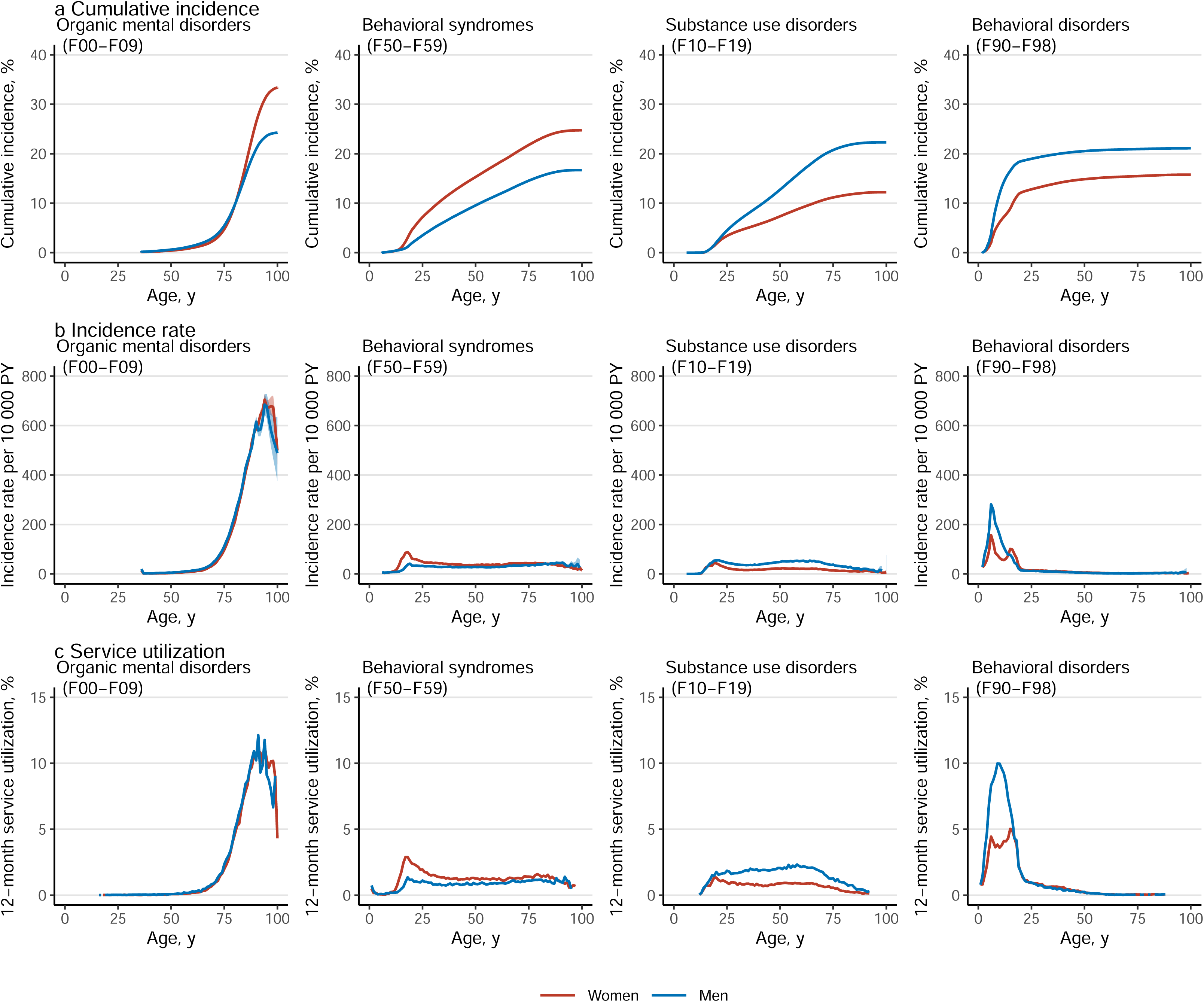
Cumulative incidence, incidence rate, and 12-month service utilization of mental disorders by gender and diagnosis ^1^ Service utilization is the number of individuals with any medical contacts with a diagnosis of a mental disorders during the year 2019, divided by the number of individuals in the study population on December 31, 2019.

**Fig. 4:**
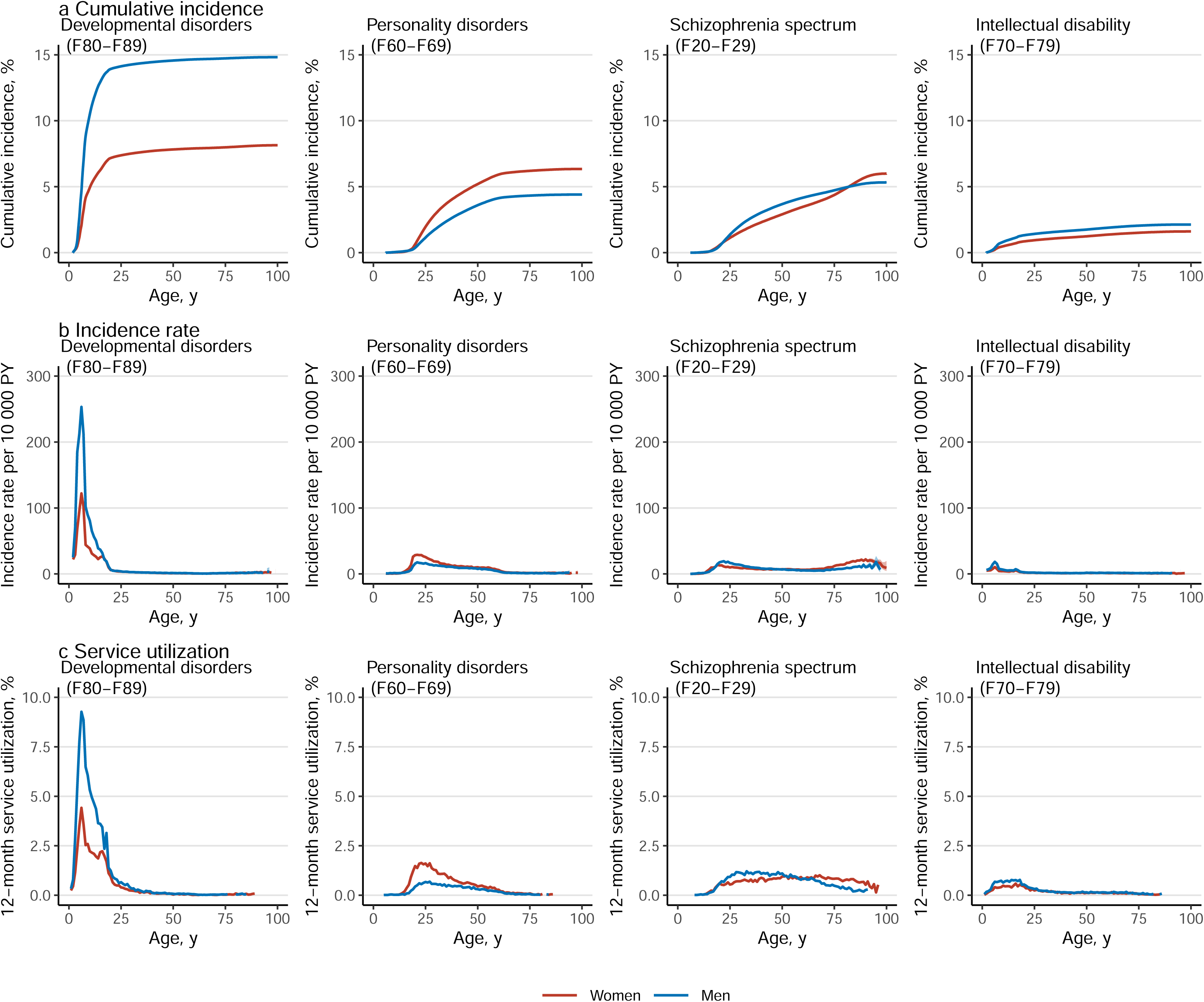
Cumulative incidence, incidence rate, and 12-month service utilization of mental disorders by gender and diagnosis ^1^ Service utilization is the number of individuals with any medical contacts with a diagnosis of a mental disorders during the year 2019, divided by the number of individuals in the study population on December 31, 2019.

After adolescence, the lowest incidence rates were observed at the age of 64 in women and 39 in men (Fig. 1c and 1d). Thereafter, the most incident disorders were dementias (F00–03), but schizophrenia spectrum (F20–F29), mood disorders (F30–F39), and behavioral syndromes (F50–F59) also showed a little increase in incidence rates at late life (Figs. 2b, 3b, and 4b).

Table 1 shows the gender-specific median age of onset and interquartile range (IQR) for different mental disorders; for non-organic mental disorders (F10–99) the median age of onset was 24.1 (interquartile range 14.8–43.3) in women and 20.0 (7.3–42.2) in men.

### 12-month Service Utilization

Overall, 9.0% of women and 7.7% of men under the age of 100 years had any medical contact with a diagnosis of a mental disorder in 2019 (Table 2). The highest service utilization, 18.1% was observed at the age of 18 in women and 16.0% in men at the age of 6 (Fig. 1e and 1f).

**Table 2:**
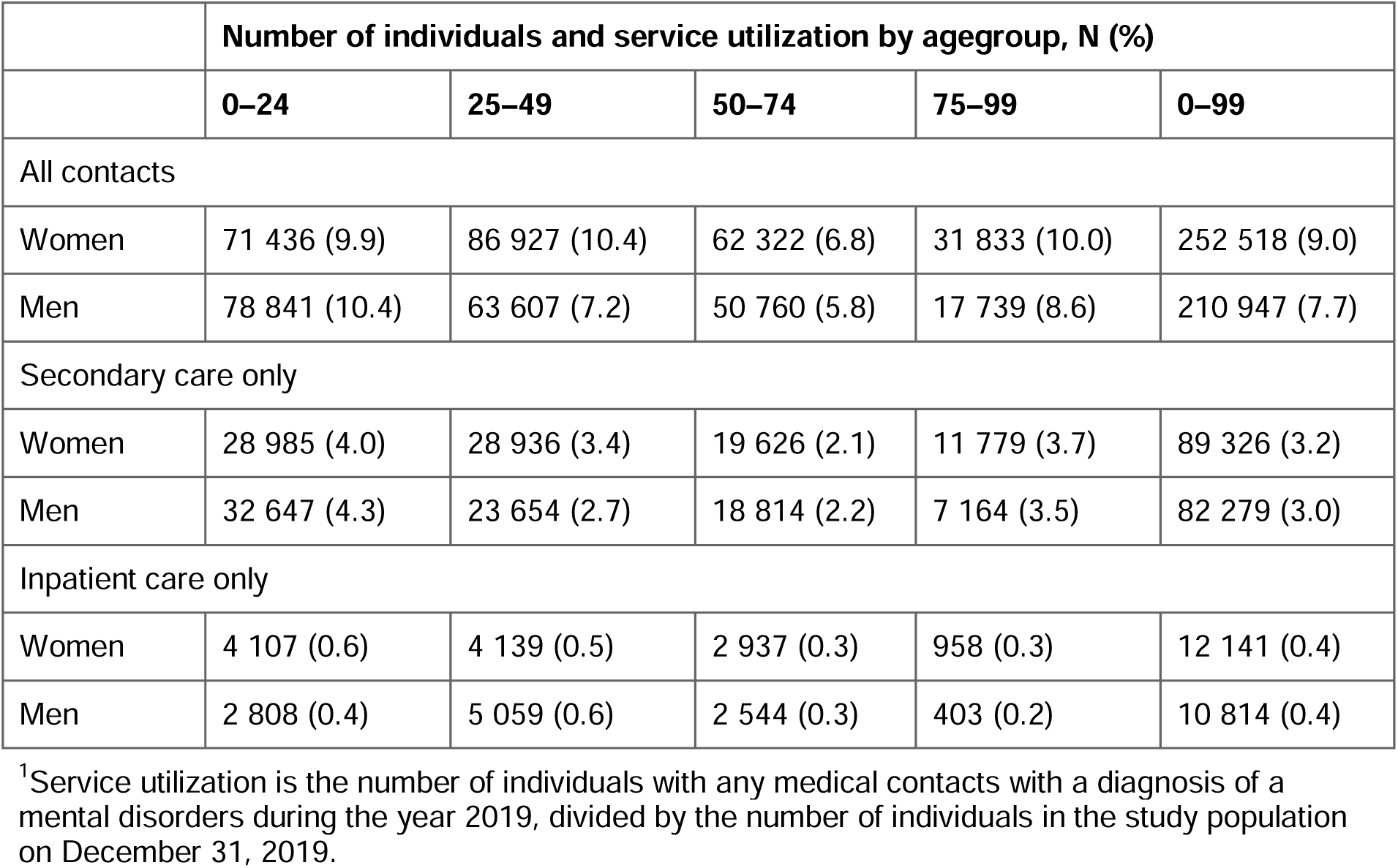
12-month service utilization for medical contacts with diagnosed mental disorders by gender, age group, and type of contact in 2019

|  | **Number of individuals and service utilization by agegroup, N (%)** |  |  |  |  |
| --- | --- | --- | --- | --- | --- |
|  | **0–24** | **25–49** | **50–74** | **75–99** | **0–99** |
| All contacts |  |  |  |  |  |
| Women | 71 436 (9.9) | 86 927 (10.4) | 62 322 (6.8) | 31 833 (10.0) | 252 518 (9.0) |
| Men | 78 841 (10.4) | 63 607 (7.2) | 50 760 (5.8) | 17 739 (8.6) | 210 947 (7.7) |
| Secondary care only |  |  |  |  |  |
| Women | 28 985 (4.0) | 28 936 (3.4) | 19 626 (2.1) | 11 779 (3.7) | 89 326 (3.2) |
| Men | 32 647 (4.3) | 23 654 (2.7) | 18 814 (2.2) | 7 164 (3.5) | 82 279 (3.0) |
| Inpatient care only |  |  |  |  |  |
| Women | 4 107 (0.6) | 4 139 (0.5) | 2 937 (0.3) | 958 (0.3) | 12 141 (0.4) |
| Men | 2 808 (0.4) | 5 059 (0.6) | 2 544 (0.3) | 403 (0.2) | 10 814 (0.4) |

<sup>1</sup>Service utilization is the number of individuals with any medical contacts with a diagnosis of a mental disorders during the year 2019, divided by the number of individuals in the study population on December 31, 2019.

Service utilization diminished throughout adulthood for most ICD–10 sub-chapter categories, with organic mental disorders (F00–F09) being an obvious exception. In addition, schizophrenia spectrum (F20–F29) and substance use disorders (F10–F19) showed relatively stable service utilization in women throughout adulthood. In men, service utilization related to substance use disorders (F10–F19) increased during adulthood, and it was the most commonly present ICD–10 sub-chapter category between ages 58 and 72 (Supplementary Table 5 and the interactive online material).

### Sensitivity analyses

In the sensitivity analysis with an additional three-year washout period for inpatient and secondary outpatient treatments and a one-year retrospective washout for primary care, a lifetime cumulative incidence of 77.7% (77.6–77.8) in women and 70.9% (70.8–71.0) in men at age 100 was observed for all disorders. In the second sensitivity analysis with increased washout periods and follow-up restricted to 2012–2020, the corresponding estimates were 86.3% (86.2–86.4) and 81.0% (80.9–81.1).

## Discussion

This nationwide cohort study with a 21-year follow-up provides comprehensive estimates of the lifetime cumulative incidence, age of onset, and 12-month service utilization for mental disorders across both primary and secondary healthcare services in Finland. Our findings indicate that 77% of women and 70% of men are affected by the age of 100, and 9.0% of women and 7.7% of men have a medical contact with a mental disorder diagnosis within a 12-month period. The highest incidence and service utilization occurred in childhood for boys and in adolescence for girls, with a second peak at around 90 years due to dementia. To our knowledge, this study provides the most extensive analysis of mental disorder incidence and service utilization throughout the life-course, with diagnosis, gender, and age-specific results.

Our estimates of lifetime cumulative incidence are lower than previous Danish estimates (87.5% for women and 76.7% for men), where medication use was used as a proxy for primary care contacts [1]. However, in children and youths, our cumulative incidence estimates are higher than those in previous Danish research, likely due to the common use of non-pharmacological treatments in this age group [1, 12]. On the other hand, our cumulative incidence estimates at age 75 are higher compared to those previously reported in WHO World Mental Helath Survey data [2]. Comprehensive lifetime risk estimates are essential for understanding the nature and impact of mental disorders. For example, excess mortality estimates are lower when all disorders, not just those treated in secondary care, are considered [19].

Schizophrenia-spectrum diagnoses exhibited a relatively high lifetime risk, with incidence persisting throughout the life course and prevalence higher in women, as expected from previous research [20]. In terms of psychiatric secondary care, current estimates of the lifetime risk for schizophrenia-spectrum disorders are lower than recent estimates from Denmark but higher than those reported approximately a decade ago [7, 8]. The lifetime risk of narrow schizophrenia (F20) was found to be lower than in the two prior Danish studies. Generally, cohort- and register-based studies have reported higher incidences of psychotic disorders compared to first-contact studies [21].

The results for specific psychotic disorders should be interpreted with caution. Unspecified psychosis had the highest lifetime incidence among schizophrenia spectrum diagnoses. However, studies focusing on specific psychotic disorders often need specialized algorithms to capture diagnoses correctly [22]. Such algorithms were not applied here as the main focus was on overall lifetime risk of any disorders. Therefore, some unspecified psychoses may later be reclassified for example as affective or substance use disorders [23]. Nevertheless, unspecified psychosis is the most common diagnosis at discharge after first hospitalization for psychosis in Finland [24], reflecting that a category of psychotic disorder without stricter criteria is practical in clinical use.

To our knowledge, this is the first study to analyze the incidence of all mental disorders using nationwide data from both primary and secondary health care. Unlike a recent meta-analysis, which mainly included studies focused on young individuals [11], our data reveal a bimodal pattern with distinct gender differences. In our study, boys showed a prominent peak at age six, associated with developmental and behavioral disorders, and a smaller peak at age 20. Girls had a smaller peak at age six but a strong peak at ages 16–18 due to mood and anxiety disorders. A Danish register study using secondary care data reported similar median age of onset estimates and also found earlier incidence peaks in boys, although the early peak at age six was absent, whereas in Sweden the early peak in boys was observed [5, 7]. In line with previous research, depression and anxiety were more common and had an earlier onset age in women compared to men. The reasons for this disparity remain unclear and are an important area for further study [25, 26].

Current 12-month service utilization estimates in young people are a few percentage points smaller than previously reported prevalence estimates [4, 15, 27]. Medical contacts with diagnoses of developmental and behavioral disorders remain prevalent during childhood and adolescence but decrease sharply by age 25. Similarly, a declining pattern was seen with respect to other diagnoses in young adulthood, suggesting a favorable course for most childhood and adolescent disorders [28], except for substance use disorders and schizophrenia spectrum disorders, which showed relatively stable patterns of service utilization through adulthood.

It is important to consider features of the Finnish healthcare system when interpreting our results. Universal health screenings are conducted in child welfare clinics, and school readiness is assessed in preschools before children begin elementary school at age seven. These screenings likely contribute to the observed peaks in neurodevelopmental and behavioral problem incidence at age six. For men, another screening occurs at age 18, before compulsory military or civil service, which may explain the increased incidence and service utilization seen in men aged 18–20 [29]. Finally, the observed incidence and the patterns of service utilization across different age groups also reflect the organization and resources of the healthcare system. For example, some changes in service utilization may be linked to transition ages in mental health services [30]. In Finland, specialized services are divided into child, adolescent, and adult psychiatry, and the transition from adolescent to adult services around age 20 may increase the risk of dropout.

This study adds comprehensive data to the body of literature showing that most individuals experience a mental disorder at some point of their life, most commonly in childhood and adolescence [1–3]. The precise definition of a correct diagnostic threshold for mental disorders is a complex question without a clear answer [31–33], and in practice, diagnoses may serve various clinical and administrative functions [34–36]. This points towards a pragmatic view on the nature of mental disorders in healthcare settings; it is possible that some of the diagnosed disorders might better be conceptualized using the broader term mental health conditions [37, 38]. Furthermore, this study clearly demonstrates the need for diagnostic systems that are usable for primary care practitioners.

The main strength of this study is its inclusion of both primary and secondary care data, because mental disorders are commonly treated in primary care [19, 39]. Universal access to publicly funded care and well-trained general practitioners likely ensures most clinically significant disorders are captured in lifetime estimates. Previous studies have shown variations in included diagnoses and age definitions for lifetime risk [1, 7, 8]. We provided estimates with and without organic mental disorders, as well as diagnosis-specific estimates up to age 100.

This study also has limitations. Ideally, the same individuals would be followed from birth to death to evaluate the lifetime risk of mental disorders. Without such data, uncertainties in lifetime risk estimates persist. To recognize incident cases, a washout period up to 25 years was utilized. Hence, likely not all prevalent disorders were recognized, particularly among the oldest individuals. Based on our sensitivity analyses, however, misclassification of prevalent cases did not appear to overestimate the cumulative incidence estimates substantially. Instead, the sensitivity analyses restricted to more recent study periods resulted in higher estimates, probably due to increased mental health treatments particularly among young people [40, 41], but this study did not aimed at examining these secular trends. On the other hand, unmet needs in psychiatric services is a well-known problem and some people may not seek treatment. It is likely that some cases are not included or they may enter with delay. Private and employer-paid mental health outpatient care are significant components of the Finnish health care system but were not comprehensively covered in the registers for the study period, and diagnostic accuracy in the registers has not been evaluated recently [17, 42, 43]. Finally, while align with previous reports from other Nordic countries, our findings may not be generalizable elsewhere, due to differences in morbidity, healthcare systems, and sociocultural circumstances.

## Conclusions

This nationwide register study with primary and secondary care data shows that most individuals experience mental disorders at some point in their lives, indicating a high need for mental health services, particularly among young people. Our findings highlight the importance of carefully selecting source populations in studies of mental disorders to ensure the generalizability of results. Future studies are needed to understand the long-term consequences of the entire spectrum of mental disorders.

## Acknowledgements

This study was funded by the European Union (ERC, MENTALNET, 101040247) and the Research Council of Finland (354237 to Dr Hakulinen; 339390 to Dr Elovainio; 352602 to Dr Pirkola).

## Disclaimer

Views and opinions expressed in this article are those of the authors only and do not necessarily reflect those of the European Union or the European Research Council.

## Contributions

Conceptualisation: Gutvilig, Hakulinen, Niemi, Suokas. Data curation: Gutvilig, Hakulinen, Niemi, Suokas. Formal analysis: Gutvilig, Niemi, Suokas. Funding acquisition: Hakulinen. Methodology: Gutvilig, Hakulinen, NcGrath, Niemi, Suokas. Resources: Elovainio, Hakulinen, Lumme, Pirkola, Suvisaari. Supervision: Hakulinen. Visualisation: Suokas. Writing – original draft: Suokas. Writing – review & editing: all authors.

## Data availability

The data that support the findings of this study are available from the National Institute of Health and Welfare (www.thl.fi) and Statistics Finland (www.stat.fi). Restrictions apply to the availability of these data, which were used under license for this study. Inquiries about secure access to data should be directed to data permit authority Findata (findata.fi/en)

## Code availability

Code used for analysis is available online at https://github.com/kmmsks/lifetime

## Conflict of interests

None reported.

## Notes

### Competing Interest Statement

The authors have declared no competing interest.

### Author Declarations

The Research Ethics Committee of the Finnish Institute for Health and Welfare approved the study protocol. Data were linked with permission from Statistics Finland and the Finnish Institute of Health and Welfare. Informed consent is not required for register-based studies in Finland.

