## Supplementary Information for "Lifetime incidence and age of onset of mental disorders, and 12-month service utilization in primary and secondary care: a Finnish nationwide registry study"

| **Supplementary Table 1:** Diagnostic classification of mental disorders according to the ICD–10, earliest possible age of onset, and the counts of cases and person-years included in the follow-up, and count of persons at risk at the start of the follow-up by gender | 2 |
| --- | --- |
| **Supplementary Fig. 1:** Example timelines in the main analysis and in sensitivity analyses with additional washout periods | 8 |
| **Supplementary Table 2:** Cumulative incidence of mental disorders in psychiatric secondary care at the ages of 25, 50, 75, and 100 years, and median age of onset (AOO) and interquartile range (IQR) by gender and ICD–10 sub-chapter category | 11 |
| **Supplementary Table 3:** Cumulative incidence of diagnosed mental disorders in psychiatric inpatient care at the ages of 25, 50, 75, and 100 years, and median age of onset (AOO) and interquartile range (IQR) by gender and ICD–10 sub-chapter category | 13 |
| **Supplementary Table 4:** Cumulative incidence of mental disorders at the ages of 25, 50, 75, and 100 years, and median age of onset (AOO) and interquartile range (IQR) by gender and diagnosis | 15 |
| **Supplementary Fig. 2:** Cumulative incidence of mental disorders by gender and ICD–10 sub-chapter category | 24 |
| **Supplementary Table 5**: 12-month service utilization for medical contacts with diagnosed mental disorders by gender, age group, and diagnosis in 2019 | 25 |

### Supplementary Table 1: Diagnostic classification of mental disorders according to the ICD–10, earliest possible age of onset, and the counts of cases and person-years included in the follow-up, and count of persons at risk at the start of the follow-up by gender

|  | | **Women** | | | **Men** | | |
| --- | --- | --- | --- | --- | --- | --- | --- |
| **Diagnosis** | **Earliest possible age of onset, y** | **No. of persons at risk at initiation of follow-up** | **Person-years** | **No. of new cases during follow-up** | **No. of persons at risk at initiation of follow-up** | **Person-years** | **No. of new cases during follow-up** |
| **Any mental disorder** | NA | 3 228 126 | 49 809 682 | 963 499 | 3 127 927 | 48 729 941 | 773 505 |
| *Non-organic mental disorders (F10–F99)* | NA | 3 243 841 | 50 427 848 | 825 651 | 3 134 567 | 49 064 737 | 686 896 |
| **Organic mental disorders (F00–F09)** | 35 | 2 279 007 | 34 042 310 | 200 581 | 2 159 837 | 31 304 083 | 132 081 |
| Dementias (F00–03) | 35 | 2 282 346 | 34 243 983 | 168 199 | 2 164 471 | 31 513 505 | 100 973 |
| Others (F04–09) | 35 | 2 298 335 | 34 753 989 | 45 756 | 2 169 023 | 31 676 455 | 41 398 |
| **Substance use disorders (F10–F19)** | 5 | 3 269 357 | 55 210 745 | 103 379 | 3 141 302 | 52 047 888 | 200 607 |
| Alcohol use disorders (F10) | 5 | 3 275 231 | 55 519 905 | 68 130 | 3 147 678 | 52 447 699 | 152 951 |
| Opiods use disorders (F11) | 5 | 3 294 587 | 56 326 049 | 5 917 | 3 216 333 | 54 463 510 | 11 164 |
| Cannabinoids use disorders (F12) | 5 | 3 294 920 | 56 345 712 | 3 542 | 3 216 108 | 54 453 801 | 11 992 |
| Sedatives or hypnotics use disorders (F13) | 5 | 3 289 392 | 56 204 896 | 9 948 | 3 211 397 | 54 360 248 | 14 901 |
| Cocaine use disorders (F14) | 5 | 3 295 246 | 56 376 168 | 104 | 3 217 508 | 54 559 274 | 271 |
| Other stimulants use disorders (F15) | 5 | 3 294 706 | 56 340 588 | 3 292 | 3 216 017 | 54 478 693 | 7 480 |
| Hallucinogens use disorders (F16) | 5 | 3 295 185 | 56 372 353 | 447 | 3 217 264 | 54 546 433 | 1 391 |
| Tobacco use disorders (F17) | 5 | 3 295 199 | 56 304 488 | 13 368 | 3 217 513 | 54 484 875 | 16 158 |
| Volatile solvents use disorders (F18) | 5 | 3 295 263 | 56 375 464 | 236 | 3 217 493 | 54 556 206 | 595 |
| Multiple and other substances use disorders (F19) | 5 | 3 294 287 | 56 286 990 | 9 623 | 3 215 158 | 54 368 320 | 20 050 |
| **Schizophrenia spectrum (F20–F29)** | 5 | 3 258 331 | 55 419 530 | 46 431 | 3 186 418 | 53 698 645 | 43 990 |
| Schizophrenia (F20) | 5 | 3 275 866 | 55 954 911 | 12 681 | 3 198 793 | 54 121 567 | 15 739 |
| Schizotypal disorder (F21) | 5 | 3 294 821 | 56 340 607 | 2 651 | 3 216 951 | 54 514 960 | 3 515 |
| Persistent delusional disorders (F22) | 5 | 3 292 016 | 56 214 872 | 16 313 | 3 215 864 | 54 457 141 | 9 721 |
| Acute and transient psychotic disorders (F23) | 5 | 3 292 743 | 56 240 916 | 9 934 | 3 215 555 | 54 445 105 | 9 381 |
| Induced delusional disorder (F24) | 5 | 3 295 237 | 56 375 877 | 115 | 3 217 565 | 54 561 262 | 88 |
| Schizoaffective disorders (F25) | 5 | 3 293 369 | 56 272 310 | 7 378 | 3 216 204 | 54 488 019 | 5 644 |
| Other nonorganic psychotic disorders (F28) | 5 | 3 295 084 | 56 358 854 | 1 826 | 3 217 418 | 54 546 653 | 1 505 |
| Unspecified nonorganic psychosis (F29) | 5 | 3 292 723 | 56 115 603 | 25 478 | 3 215 203 | 54 303 683 | 27 129 |
| **Mood disorders (F30–F39)** | 5 | 3 233 574 | 52 507 527 | 370 659 | 3 172 870 | 52 111 491 | 227 796 |
| Mania and bipolar disorder (F30–F31) | 5 | 3 290 388 | 56 007 834 | 33 420 | 3 213 431 | 54 284 500 | 25 220 |
| Depressive disorders (F32–F33) | 5 | 3 239 076 | 52 753 433 | 353 270 | 3 177 500 | 52 323 031 | 211 412 |
| Others (F34–F39) | 5 | 3 292 186 | 55 971 491 | 48 964 | 3 215 785 | 54 327 029 | 30 187 |
| **Neurotic, stress–related and somatoform disorders (F40–F48)** | 5 | 3 250 441 | 52 573 517 | 476 006 | 3 172 282 | 52 088 711 | 251 879 |
| Anxiety disorders (F40–F41) | 5 | 3 279 434 | 54 463 840 | 265 500 | 3 201 642 | 53 315 011 | 150 218 |
| Obsessive–compulsive disorder (F42) | 5 | 3 294 220 | 56 249 750 | 16 551 | 3 216 372 | 54 456 139 | 11 821 |
| Reaction to severe stress, and adjustment disorders (F43) | 5 | 3 289 329 | 55 081 989 | 178 647 | 3 210 584 | 53 824 726 | 83 726 |
| Dissociative disorders (F44) | 5 | 3 294 917 | 56 324 996 | 6 405 | 3 217 423 | 54 544 173 | 2 011 |
| Somatoform disorders (F45) | 5 | 3 292 821 | 55 846 137 | 91 633 | 3 216 293 | 54 326 902 | 37 740 |
| Other neurotic disorders (F48) | 5 | 3 294 823 | 56 341 437 | 3 252 | 3 217 313 | 54 541 732 | 1 829 |
| **Behavioral syndromes (F50–F59)** | 5 | 3 287 922 | 55 108 211 | 222 534 | 3 212 080 | 53 785 916 | 147 841 |
| Eating disorders (F50) | 5 | 3 291 390 | 56 059 227 | 28 846 | 3 217 003 | 54 524 081 | 3 580 |
| Nonorganic sleep disorders (F51) | 5 | 3 293 061 | 55 732 016 | 137 323 | 3 213 391 | 54 082 350 | 88 422 |
| Disorders associated with the puerperium (F53) | 5 | 3 294 469 | 56 340 927 | 2 323 | 3 217 576 | 54 561 752 | 52 |
| Other (F54-F59) | 5 | 3 295 076 | 56 341 279 | 4 250 | 3 217 276 | 54 479 986 | 15 839 |
| **Personality disorders (F60–F69)** | 5 | 3 278 518 | 55 640 558 | 52 725 | 3 190 990 | 53 768 851 | 38 016 |
| Paranoid personality disorder (F60.0) | 5 | 3 294 727 | 56 349 096 | 2 015 | 3 216 580 | 54 528 648 | 1 874 |
| Schizoid personality disorder (F60.1) | 5 | 3 294 802 | 56 358 688 | 1 026 | 3 216 158 | 54 518 663 | 1 930 |
| Dissocial personality disorder (F60.2) | 5 | 3 294 968 | 56 366 623 | 673 | 3 214 692 | 54 488 729 | 3 040 |
| Emotionally unstable personality disorder (F60.3) | 5 | 3 292 241 | 56 129 717 | 23 077 | 3 214 908 | 54 439 531 | 8 458 |
| Histrionic personality disorder (F60.4) | 5 | 3 294 792 | 56 365 191 | 381 | 3 217 362 | 54 557 441 | 149 |
| Anankastic personality disorder (F60.5) | 5 | 3 295 007 | 56 325 286 | 6 359 | 3 217 174 | 54 527 217 | 3 309 |
| Anxious [avoidant] personality disorder (F60.6) | 5 | 3 295 034 | 56 349 608 | 3 125 | 3 217 148 | 54 529 442 | 3 148 |
| Dependent personality disorder (F60.7) | 5 | 3 293 814 | 56 325 792 | 2 294 | 3 216 501 | 54 535 728 | 784 |
| Other specific personality disorders (F60.8) | 5 | 3 290 627 | 56 262 909 | 3 768 | 3 210 182 | 54 401 996 | 3 551 |
| Personality disorder, unspecified (F60.9) | 5 | 3 291 156 | 56 216 515 | 9 662 | 3 209 427 | 54 353 438 | 7 455 |
| Mixed and other personality disorders (F61) | 5 | 3 293 635 | 56 228 979 | 12 584 | 3 215 143 | 54 412 088 | 10 831 |
| Enduring personality changes, not attributable to brain damage and disease (F62) | 5 | 3 295 217 | 56 371 958 | 553 | 3 217 509 | 54 556 620 | 556 |
| Others (F63–F69) | 5 | 3 294 654 | 56 329 151 | 6 182 | 3 215 917 | 54 482 560 | 7 503 |
| **Intellectual disability (F70–F79)** | 1 | 3 388 475 | 58 623 088 | 12 139 | 3 314 325 | 56 875 882 | 16 521 |
| Mild intellectual disability (F70) | 1 | 3 393 240 | 58 746 569 | 6 772 | 3 319 293 | 57 019 905 | 8 948 |
| Moderate intellectual disability (F71) | 1 | 3 396 151 | 58 829 427 | 2 689 | 3 323 099 | 57 127 928 | 3 658 |
| Severe intellectual disability (F72) | 1 | 3 397 216 | 58 861 296 | 1 369 | 3 324 080 | 57 163 625 | 1 861 |
| Profound intellectual disability (F73) | 1 | 3 397 521 | 58 870 163 | 730 | 3 324 533 | 57 178 054 | 846 |
| Other intellectual disability (F78) | 1 | 3 397 859 | 58 876 024 | 441 | 3 324 860 | 57 182 738 | 547 |
| Unspecified intellectual disability (F79) | 1 | 3 395 277 | 58 802 186 | 4 199 | 3 322 338 | 57 093 590 | 6 442 |
| **Disorders of psychological development (F80–F89)** | 1 | 3 392 069 | 58 345 673 | 57 741 | 3 310 127 | 56 006 892 | 109 259 |
| Specific developmental disorders of speech and language (F80) | 1 | 3 394 772 | 58 655 011 | 19 871 | 3 317 122 | 56 659 611 | 42 324 |
| Specific developmental disorders of scholastic skills (F81) | 1 | 3 396 610 | 58 738 952 | 15 036 | 3 321 575 | 56 897 387 | 26 335 |
| Specific developmental disorder of motor function (F82) | 1 | 3 397 453 | 58 826 458 | 4 544 | 3 323 618 | 57 035 225 | 12 733 |
| Mixed specific developmental disorders (F83) | 1 | 3 397 084 | 58 780 820 | 9 270 | 3 322 763 | 56 950 196 | 20 969 |
| Pervasive developmental disorders (F84) | 1 | 3 397 372 | 58 821 813 | 7 353 | 3 323 263 | 57 006 791 | 19 312 |
| Other disorders of psychological development (F88) | 1 | 3 397 831 | 58 868 988 | 1 676 | 3 324 776 | 57 155 120 | 4 779 |
| Unspecified disorder of psychological development (F89) | 1 | 3 397 825 | 58 871 605 | 1 091 | 3 324 806 | 57 170 084 | 2 591 |
| **Behavioral and emotional disorders (F90–F98)** | 1 | 3 389 713 | 57 874 677 | 120 914 | 3 312 032 | 55 756 858 | 165 584 |
| ADHD (F90) | 1 | 3 397 051 | 58 739 843 | 27 225 | 3 321 731 | 56 744 817 | 60 820 |
| Conduct disorders (F91) | 1 | 3 396 923 | 58 811 569 | 5 778 | 3 322 668 | 57 033 661 | 13 419 |
| Mixed disorders of conduct and emotions (F92) | 1 | 3 397 248 | 58 786 187 | 8 466 | 3 323 535 | 56 995 691 | 17 425 |
| Emotional disorders with onset specific to childhood (F93) | 1 | 3 397 301 | 58 696 754 | 23 519 | 3 324 183 | 57 017 966 | 21 124 |
| Disorders of social functioning with onset specific to childhood and adolescence (F94) | 1 | 3 397 643 | 58 843 246 | 3 879 | 3 324 572 | 57 138 753 | 5 088 |
| Tic disorders (F95) | 1 | 3 397 586 | 58 851 794 | 3 099 | 3 324 125 | 57 107 034 | 8 403 |
| Other behavioral and emotional disorders with onset usually occurring in childhood and adolescence (F98) | 1 | 3 393 821 | 58 603 186 | 24 037 | 3 319 968 | 56 807 724 | 35 776 |

### Supplementary Fig. 1: Example timelines in the main analysis and in sensitivity analyses with additional washout periods

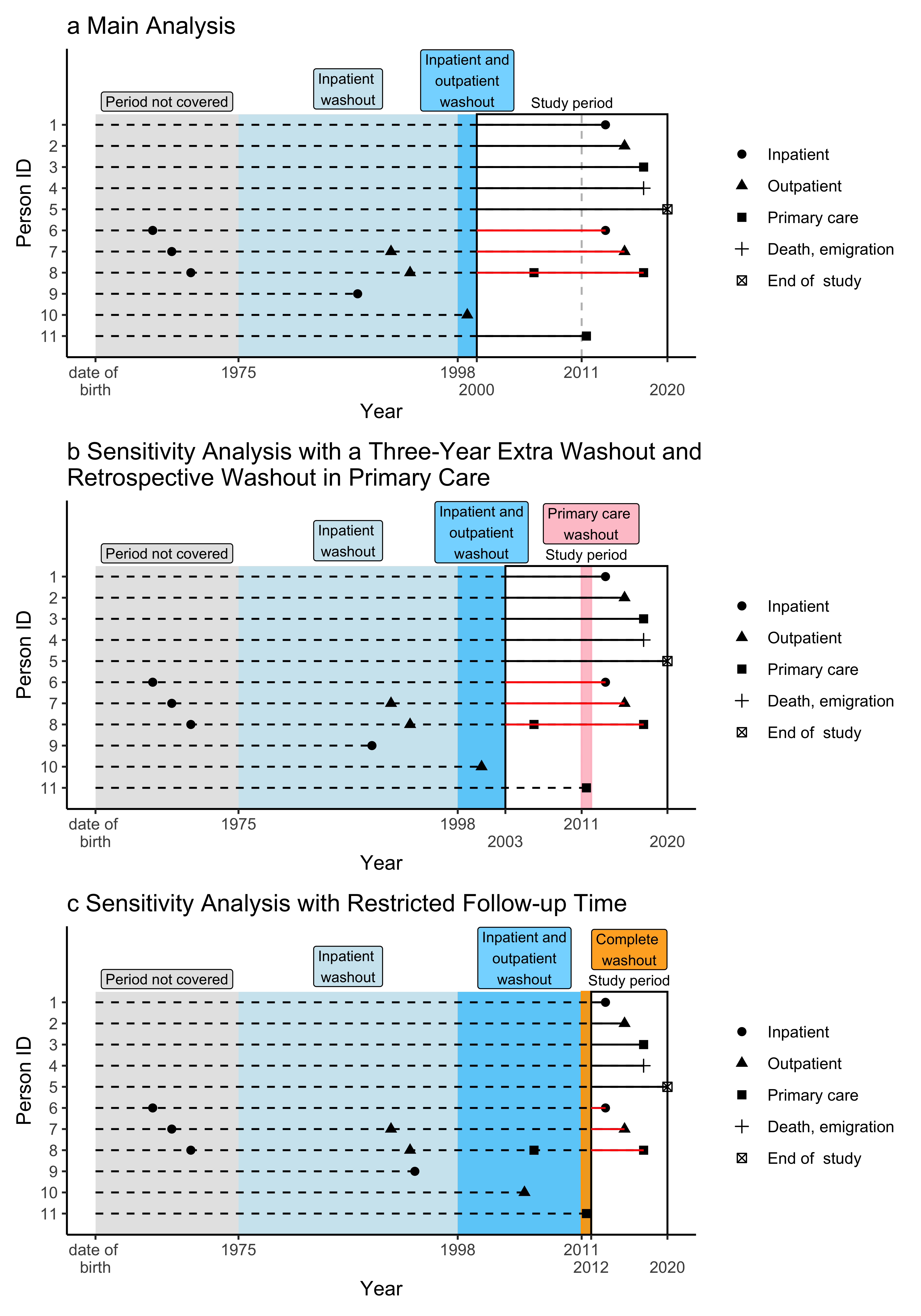

**The procedures to differentiate incident cases from prevalent cases demonstrated.** Solid black lines indicate person-time in the study at risk for truly incident cases. Red lines indicate person-time in the study where participants had previous treatments before the start of the washout period and were incorrectly included. Dashed lines indicate person-time that is not included in the study. Complete washout indicates the washout period including inpatient, outpatient and primary care treatments simultaneously.

**Panel A** demonstrates the main analysis, in which we excluded disorder-specific prevalent cases at the start of the follow-up period. This exclusion included individuals with inpatient treatments between 1975 and 1999, as well as those with outpatient care between 1998 and 1999.

**Panel B** demonstrates the sensitivity analysis with an additional three-year washout period and a retrospective washout for primary care. In this analysis, we shortened the study period from 2000–2020 to 2003–2020, incorporating an extra one-year washout for primary care. This approach aligns the washout procedure with that of a previous Danish study, where medication use served as a proxy for primary care contacts[1]. While this method is stricter in excluding prevalent cases, it introduces immortal time bias. In the Danish study, estimates of lifetime risk for mental disorders with additional 5- and 10-year washout periods were reported, with the estimates decreasing by only 0.6 and 2.0 percentage points, respectively.

**Panel C** demonstrates the sensitivity analysis with a further restricted follow-up period. In this analysis, we shortened the study period from2000–2020 to 2012–2020. The washout period for inpatient care is 1975–2011, for outpatient secondary care 1998–2011, and for primary care the year 2011. This scenario avoids introducing immortal time bias but limits the overall time span captured in the study.

The following example timelines are presented:

Person 1 has his/her first inpatient admission during the study period, with no prior outpatient or primary care mental health treatments, and is therefore correctly identified as an incident case.

Person 2 has his/her first psychiatric outpatient contact during the study period, with no prior inpatient or primary care mental health treatments, and is likewise correctly identified as an incident case.

Person 3 has his/her first contact with primary care mental health services in 2011 or later, with no prior inpatient or secondary care mental health treatments, and is thus correctly identified as an incident case.

Person 4 contributes to the risk of experiencing a mental disorder until he/she dies or emigrates from Finland.

Person 5 contributes to the risk of experiencing a mental disorder until the end of the study period.

Person 6 had his/her first inpatient admission before the launch of inpatient data in the register (1975). Consequently, he/she enters the study incorrectly and contributes to the risk of an incident mental disorder. In this case, he/she has an inpatient admission during the study period that is misclassified as an incident event.

Person 7 had his/her first inpatient admission before the launch of inpatient data in the register (1975) and a psychiatric secondary care outpatient treatment before the launch of outpatient data in the register (1998). Thus, he/she enters the study incorrectly and contributes to the risk of an incident mental disorder. In this example, he/she has an outpatient contact during the study period that is misclassified as an incident event.

Person 8 had his/her first inpatient admission before the launch of inpatient data in the register (1975), a psychiatric secondary care outpatient treatment before the launch of outpatient data in the register (1998), and a primary care mental health contact before the launch of primary care data in the register (2011). Therefore, he/she enters the study incorrectly and contributes to the risk of an incident mental disorder. In this case, he/she has a primary care mental health contact in 2011 or later and is misclassified as an incident event.

Persons 9 and 10 are examples of individuals who had inpatient care between 1975 and 1999 or secondary outpatient care between 1998 and 1999, and are correctly excluded from the study population.

Person 11 has his/her first contact with primary care mental health services in 2011 and demonstrates the additional primary care washout in the sensitivity analyses.

### Supplementary Table 2: Cumulative incidence of mental disorders in psychiatric secondary care at the ages of 25, 50, 75, and 100 years, and median age of onset (AOO) and interquartile range (IQR) by gender and ICD–10 sub-chapter category

|  | **Cumulative incidence at given ages (95% CI), %** | | | | | | | | | | **Gender ratio^1^** |
| --- | --- | --- | --- | --- | --- | --- | --- | --- | --- | --- | --- |
|  | **Women** | | | | | **Men** | | | | |  |
| **Diagnosis** | **25** | **50** | **75** | **100** | **Median AOO (IQR)** | **25** | **50** | **75** | **100** | **Median AOO (IQR)** |  |
| Any mental disorder | 18.9 (18.8–19.0) | 31.4 (31.3–31.5) | 37.3 (37.2–37.4) | 39.7 (39.6–39.8) | 26.3 (16.6–45.7) | 15.3 (15.2–15.4) | 25.1 (25.0–25.1) | 29.9 (29.8–30.0) | 31.5 (31.4–31.6) | 25.8 (14.7–45.6) | 1.26 |
| *Non-organic mental disorders (F10–F99)* | 18.9 (18.8–19.0) | 31.4 (31.3–31.5) | 37.0 (36.9–37.1) | 38.6 (38.5–38.7) | 25.6 (16.5–43.7) | 15.3 (15.2–15.4) | 25.0 (24.9–25.1) | 29.5 (29.4–29.6) | 30.4 (30.3–30.5) | 24.8 (14.3–43.2) | 1.27 |
| Organic mental disorders (F00–F09) | NA | 0.2 (0.2–0.2) | 1.2 (1.2–1.2) | 3.3 (3.2–3.3) | 78.5 (70.5–84.5) | NA | 0.3 (0.3–0.3) | 1.4 (1.4–1.5) | 2.8 (2.8–2.9) | 74.7 (62.9–81.8) | 1.16 |
| Substance use disorders (F10–F19) | 1.4 (1.4–1.5) | 3.2 (3.2–3.3) | 4.2 (4.2–4.3) | 4.3 (4.3–4.4) | 34.7 (22.0–49.8) | 1.9 (1.8–1.9) | 5.7 (5.6–5.7) | 7.4 (7.4–7.5) | 7.5 (7.5–7.6) | 36.6 (25.1–49.9) | 0.57 |
| Schizophrenia spectrum (F20–F29) | 1.1 (1.1–1.2) | 2.8 (2.7–2.8) | 4.0 (3.9–4.0) | 4.8 (4.7–4.8) | 43.0 (25.6–67.3) | 1.4 (1.4–1.5) | 3.5 (3.4–3.5) | 4.2 (4.2–4.3) | 4.5 (4.5–4.6) | 31.8 (23.0–48.3) | 1.05 |
| Mood disorders (F30–F39) | 10.8 (10.8–10.9) | 20.8 (20.7–20.9) | 25.5 (25.4–25.6) | 26.5 (26.4–26.6) | 29.8 (18.9–47.1) | 5.4 (5.4–5.5) | 12.7 (12.6–12.8) | 16.1 (16.1–16.2) | 16.7 (16.6–16.8) | 34.0 (21.8–49.4) | 1.58 |
| Neurotic, stress–related and somatoform disorders (F40–F48) | 10.5 (10.4–10.5) | 18.7 (18.7–18.8) | 21.9 (21.8–21.9) | 22.5 (22.4–22.6) | 26.6 (17.7–42.0) | 5.7 (5.7–5.8) | 10.9 (10.9–11.0) | 12.8 (12.7–12.8) | 13.1 (13.0–13.1) | 27.7 (19.1–42.6) | 1.72 |
| Behavioral syndromes (F50–F59) | 2.8 (2.8–2.9) | 4.2 (4.1–4.2) | 4.6 (4.6–4.7) | 4.7 (4.7–4.8) | 21.0 (16.1–34.2) | 0.6 (0.6–0.6) | 1.2 (1.2–1.2) | 1.5 (1.5–1.6) | 1.6 (1.6–1.6) | 32.1 (18.4–49.4) | 2.92 |
| Personality disorders (F60–F69) | 1.9 (1.9–1.9) | 4.5 (4.5–4.6) | 5.2 (5.2–5.3) | 5.3 (5.2–5.3) | 29.6 (22.3–42.4) | 1.0 (1.0–1.1) | 3.0 (3.0–3.1) | 3.6 (3.5–3.6) | 3.6 (3.5–3.6) | 32.3 (23.9–44.0) | 1.47 |
| Intellectual disability (F70–F79) | 0.2 (0.2–0.2) | 0.3 (0.3–0.3) | 0.4 (0.4–0.4) | 0.4 (0.4–0.4) | 24.3 (16.0–41.5) | 0.3 (0.3–0.3) | 0.4 (0.4–0.4) | 0.5 (0.5–0.5) | 0.5 (0.5–0.5) | 20.9 (13.5–36.4) | 0.84 |
| Disorders of psychological development (F80–F89) | 1.6 (1.5–1.6) | 1.8 (1.8–1.8) | 1.8 (1.8–1.8) | 1.8 (1.8–1.8) | 14.0 (9.7–18.0) | 3.4 (3.4–3.5) | 3.7 (3.7–3.8) | 3.7 (3.7–3.8) | 3.7 (3.7–3.8) | 10.8 (8.0–15.0) | 0.48 |
| Behavioral and emotional disorders (F90–F98) | 6.0 (5.9–6.0) | 7.4 (7.3–7.4) | 7.8 (7.7–7.9) | 7.9 (7.8–7.9) | 15.7 (11.9–24.2) | 7.8 (7.8–7.9) | 9.0 (8.9–9.0) | 9.2 (9.2–9.3) | 9.3 (9.2–9.3) | 11.7 (8.5–17.0) | 0.85 |
| *^1^Women to men ratio of the cumulative incidence estimates at the age of 100.* | | | | | | | | | | | |

### Supplementary Table 3: Cumulative incidence of diagnosed mental disorders in psychiatric inpatient care at the ages of 25, 50, 75, and 100 years, and median age of onset (AOO) and interquartile range (IQR) by gender and ICD–10 sub-chapter category

|  | **Cumulative incidence at given ages (95% CI), %** | | | | | | | | | | **Gender ratio^1^** |
| --- | --- | --- | --- | --- | --- | --- | --- | --- | --- | --- | --- |
|  | **Women** | | | | | **Men** | | | | |  |
| **Diagnosis** | **25** | **50** | **75** | **100** | **Median AOO (IQR)** | **25** | **50** | **75** | **100** | **Median AOO (IQR)** |  |
| Any mental disorder | 5.3 (5.3–5.4) | 9.1 (9.0–9.1) | 11.5 (11.5–11.6) | 13.3 (13.3–13.4) | 32.2 (18.1–58.4) | 4.6 (4.6–4.7) | 8.9 (8.8–8.9) | 11.3 (11.2–11.4) | 12.5 (12.4–12.5) | 33.0 (19.7–53.5) | 1.07 |
| *Non-organic mental disorders (F10–F99)* | 5.3 (5.3–5.4) | 9.1 (9.0–9.1) | 11.3 (11.2–11.3) | 12.3 (12.2–12.4) | 29.1 (17.5–51.4) | 4.6 (4.6–4.7) | 8.8 (8.8–8.9) | 10.9 (10.9–11.0) | 11.4 (11.4–11.5) | 30.2 (19.0–48.2) | 1.08 |
| Organic mental disorders (F00–F09) | NA | 0.1 (0.1–0.1) | 0.7 (0.7–0.7) | 2.0 (2.0–2.0) | 78.9 (72.0–84.5) | NA | 0.1 (0.1–0.1) | 0.8 (0.8–0.8) | 1.8 (1.8–1.8) | 76.2 (66.7–82.6) | 1.10 |
| Substance use disorders (F10–F19) | 0.7 (0.7–0.7) | 1.7 (1.7–1.7) | 2.2 (2.2–2.3) | 2.3 (2.2–2.3) | 35.2 (22.5–50.1) | 1.1 (1.0–1.1) | 3.4 (3.3–3.4) | 4.4 (4.4–4.5) | 4.5 (4.4–4.5) | 36.9 (25.6–50.0) | 0.51 |
| Schizophrenia spectrum (F20–F29) | 0.8 (0.8–0.8) | 2.0 (2.0–2.0) | 2.9 (2.8–2.9) | 3.4 (3.4–3.5) | 43.0 (26.1–67.5) | 1.1 (1.0–1.1) | 2.6 (2.6–2.6) | 3.2 (3.1–3.2) | 3.4 (3.3–3.4) | 31.5 (23.2–47.6) | 1.02 |
| Mood disorders (F30–F39) | 3.2 (3.2–3.2) | 5.9 (5.8–5.9) | 7.4 (7.3–7.5) | 7.9 (7.9–8.0) | 30.7 (18.8–50.6) | 1.6 (1.6–1.6) | 4.0 (4.0–4.1) | 5.3 (5.3–5.4) | 5.6 (5.5–5.6) | 36.9 (23.4–51.7) | 1.42 |
| Neurotic, stress–related and somatoform disorders (F40–F48) | 2.0 (2.0–2.0) | 3.4 (3.3–3.4) | 4.0 (3.9–4.0) | 4.2 (4.1–4.2) | 25.8 (17.4–43.3) | 1.2 (1.2–1.3) | 2.4 (2.4–2.4) | 2.8 (2.7–2.8) | 2.8 (2.8–2.9) | 27.8 (19.6–42.2) | 1.47 |
| Behavioral syndromes (F50–F59) | 0.7 (0.7–0.7) | 0.9 (0.9–1.0) | 1.0 (1.0–1.0) | 1.0 (1.0–1.0) | 19.3 (15.7–29.0) | 0.1 (0.1–0.1) | 0.2 (0.2–0.2) | 0.2 (0.2–0.2) | 0.2 (0.2–0.2) | 34.8 (20.8–51.6) | 4.50 |
| Personality disorders (F60–F69) | 0.7 (0.7–0.7) | 1.6 (1.5–1.6) | 1.8 (1.8–1.8) | 1.8 (1.8–1.9) | 29.3 (22.2–41.9) | 0.3 (0.3–0.4) | 1.1 (1.1–1.2) | 1.3 (1.3–1.3) | 1.3 (1.3–1.3) | 33.0 (24.6–43.7) | 1.38 |
| Intellectual disability (F70–F79) | 0.1 (0.1–0.1) | 0.1 (0.1–0.2) | 0.2 (0.2–0.2) | 0.2 (0.2–0.2) | 27.6 (17.7–43.9) | 0.1 (0.1–0.1) | 0.2 (0.2–0.2) | 0.2 (0.2–0.2) | 0.2 (0.2–0.2) | 26.1 (17.4–43.1) | 0.86 |
| Disorders of psychological development (F80–F89) | 0.3 (0.3–0.3) | 0.4 (0.3–0.4) | 0.4 (0.3–0.4) | 0.4 (0.3–0.4) | 14.8 (12.1–17.7) | 0.7 (0.7–0.7) | 0.7 (0.7–0.8) | 0.7 (0.7–0.8) | 0.7 (0.7–0.8) | 12.1 (9.4–16.2) | 0.48 |
| Behavioral and emotional disorders (F90–F98) | 1.4 (1.4–1.4) | 1.4 (1.4–1.5) | 1.5 (1.4–1.5) | 1.5 (1.4–1.5) | 14.8 (12.9–16.5) | 1.8 (1.8–1.8) | 1.9 (1.9–1.9) | 1.9 (1.9–2.0) | 1.9 (1.9–2.0) | 11.9 (9.4–15.3) | 0.76 |
| *^1^Women to men ratio of the cumulative incidence estimates at the age of 100.* | | | | | | | | | | | |

### Supplementary Table 4: Cumulative incidence of mental disorders at the ages of 25, 50, 75, and 100 years, and median age of onset (AOO) and interquartile range (IQR) by gender and diagnosis

|  | **Cumulative incidence at given ages (95% CI), %** | | | | | | | | | | **Gender ratio^1^** |
| --- | --- | --- | --- | --- | --- | --- | --- | --- | --- | --- | --- |
|  | **Women** | | | | | **Men** | | | | |  |
| **Diagnosis** | **25** | **50** | **75** | **100** | **Median AOO (IQR)** | **25** | **50** | **75** | **100** | **Median AOO (IQR)** |  |
| **Any mental disorder** | 35.8 (35.7–35.9) | 56.0 (55.9–56.1) | 66.6 (66.5–66.6) | 76.7 (76.6–76.7) | 27.2 (15.5–52.7) | 36.3 (36.2–36.4) | 51.0 (50.9–51.1) | 61.3 (61.2–61.4) | 69.7 (69.6–69.8) | 23.4 (8.2–52.7) | 1.10 |
| *Non-organic mental disorders (F10–F99)* | 35.8 (35.7–35.9) | 56.0 (55.9–56.1) | 65.6 (65.5–65.7) | 69.3 (69.2–69.4) | 24.1 (14.8–43.3) | 36.3 (36.2–36.4) | 50.9 (50.8–51.0) | 60.0 (59.9–60.1) | 62.7 (62.6–62.8) | 20.0 (7.3–42.2) | 1.10 |
| **Organic mental disorders (F00–F09)** | NA | 0.5 (0.4–0.5) | 5.5 (5.4–5.5) | 33.4 (33.3–33.6) | 83.5 (77.9–88.2) | NA | 0.7 (0.6–0.7) | 6.1 (6.1–6.2) | 24.3 (24.2–24.4) | 81.1 (74.9–86.2) | 1.38 |
| Dementias (F00–03) | NA | 0.1 (0.1–0.1) | 3.8 (3.7–3.8) | 29.0 (28.9–29.1) | 84.1 (78.9–88.6) | NA | 0.1 (0.1–0.1) | 3.7 (3.7–3.8) | 19.8 (19.7–19.9) | 82.1 (76.8–86.9) | 1.46 |
| Others (F04–09) | NA | 0.3 (0.3–0.4) | 2.1 (2.1–2.1) | 7.3 (7.3–7.4) | 81.3 (73.6–86.9) | NA | 0.6 (0.5–0.6) | 2.9 (2.8–2.9) | 6.8 (6.8–6.9) | 77.6 (67.0–84.2) | 1.08 |
| **Substance use disorders (F10–F19)** | 3.6 (3.6–3.7) | 7.5 (7.5–7.6) | 11.3 (11.2–11.3) | 12.2 (12.2–12.3) | 41.6 (22.7–59.7) | 4.9 (4.8–4.9) | 13.1 (13.0–13.2) | 20.9 (20.8–21.0) | 22.3 (22.2–22.4) | 44.5 (26.8–59.9) | 0.55 |
| Alcohol use disorders (F10) | 2.3 (2.3–2.4) | 5.0 (5.0–5.1) | 7.6 (7.5–7.7) | 8.1 (8.1–8.2) | 42.4 (23.0–58.2) | 2.9 (2.8–2.9) | 9.4 (9.4–9.5) | 16.2 (16.1–16.3) | 17.3 (17.2–17.4) | 47.6 (31.1–61.0) | 0.47 |
| Opiods use disorders (F11) | 0.3 (0.2–0.3) | 0.6 (0.6–0.6) | 0.7 (0.7–0.7) | 0.7 (0.7–0.8) | 29.4 (23.2–41.8) | 0.3 (0.3–0.3) | 1.2 (1.1–1.2) | 1.3 (1.3–1.3) | 1.3 (1.3–1.3) | 30.7 (24.9–39.2) | 0.57 |
| Cannabinoids use disorders (F12) | 0.3 (0.3–0.3) | 0.4 (0.4–0.5) | 0.5 (0.4–0.5) | 0.5 (0.4–0.5) | 21.8 (18.5–27.0) | 0.8 (0.8–0.8) | 1.4 (1.4–1.4) | 1.4 (1.4–1.5) | 1.4 (1.4–1.5) | 23.5 (19.6–29.9) | 0.32 |
| Sedatives or hypnotics use disorders (F13) | 0.2 (0.2–0.2) | 0.7 (0.7–0.7) | 1.1 (1.1–1.1) | 1.2 (1.2–1.2) | 43.8 (28.1–62.7) | 0.4 (0.4–0.4) | 1.4 (1.3–1.4) | 1.7 (1.6–1.7) | 1.8 (1.7–1.8) | 34.4 (26.2–48.0) | 0.70 |
| Cocaine use disorders (F14) | 0.0 (0.0–0.0) | 0.0 (0.0–0.0) | 0.0 (0.0–0.0) | 0.0 (0.0–0.0) | 26.0 (22.0–45.0) | 0.0 (0.0–0.0) | 0.0 (0.0–0.0) | 0.0 (0.0–0.0) | 0.0 (0.0–0.0) | 30.9 (24.6–40.5) | 0.41 |
| Other stimulants use disorders (F15) | 0.2 (0.2–0.2) | 0.4 (0.4–0.4) | 0.4 (0.4–0.4) | 0.4 (0.4–0.4) | 24.3 (20.4–30.3) | 0.3 (0.3–0.4) | 0.9 (0.8–0.9) | 0.9 (0.9–0.9) | 0.9 (0.9–0.9) | 27.3 (22.6–34.0) | 0.47 |
| Hallucinogens use disorders (F16) | 0.0 (0.0–0.0) | 0.1 (0.0–0.1) | 0.1 (0.1–0.1) | 0.1 (0.1–0.1) | 23.0 (19.5–30.0) | 0.1 (0.1–0.1) | 0.2 (0.2–0.2) | 0.2 (0.2–0.2) | 0.2 (0.2–0.2) | 25.0 (20.6–32.0) | 0.34 |
| Tobacco use disorders (F17) | 0.2 (0.1–0.2) | 0.7 (0.7–0.7) | 1.5 (1.5–1.5) | 1.6 (1.5–1.6) | 52.6 (35.9–63.8) | 0.1 (0.1–0.1) | 0.7 (0.6–0.7) | 1.7 (1.7–1.8) | 1.9 (1.8–1.9) | 56.9 (43.6–66.2) | 0.85 |
| Volatile solvents use disorders (F18) | 0.0 (0.0–0.0) | 0.0 (0.0–0.0) | 0.0 (0.0–0.0) | 0.0 (0.0–0.0) | 27.3 (22.2–40.5) | 0.0 (0.0–0.0) | 0.1 (0.1–0.1) | 0.1 (0.1–0.1) | 0.1 (0.1–0.1) | 31.4 (22.8–46.7) | 0.43 |
| Multiple and other substances use disorders (F19) | 0.6 (0.6–0.6) | 1.0 (1.0–1.1) | 1.2 (1.1–1.2) | 1.2 (1.2–1.2) | 25.9 (20.2–38.5) | 1.0 (0.9–1.0) | 2.2 (2.1–2.2) | 2.3 (2.3–2.4) | 2.4 (2.3–2.4) | 27.2 (21.8–35.8) | 0.51 |
| **Schizophrenia spectrum (F20–F29)** | 1.2 (1.2–1.2) | 3.0 (2.9–3.0) | 4.5 (4.4–4.5) | 6.0 (5.9–6.0) | 50.5 (28.1–75.4) | 1.5 (1.5–1.6) | 3.7 (3.7–3.8) | 4.8 (4.7–4.8) | 5.3 (5.3–5.4) | 34.5 (23.8–55.0) | 1.13 |
| Schizophrenia (F20) | 0.3 (0.2–0.3) | 0.9 (0.9–0.9) | 1.4 (1.3–1.4) | 1.6 (1.6–1.6) | 46.1 (29.8–65.9) | 0.4 (0.4–0.5) | 1.4 (1.4–1.4) | 1.8 (1.7–1.8) | 1.9 (1.8–1.9) | 33.7 (25.3–49.3) | 0.85 |
| Schizotypal disorder (F21) | 0.1 (0.1–0.1) | 0.2 (0.2–0.2) | 0.3 (0.3–0.3) | 0.3 (0.3–0.3) | 39.4 (27.0–54.1) | 0.1 (0.1–0.1) | 0.3 (0.3–0.3) | 0.4 (0.4–0.4) | 0.4 (0.4–0.4) | 33.8 (25.2–46.2) | 0.79 |
| Persistent delusional disorders (F22) | 0.1 (0.0–0.1) | 0.4 (0.4–0.4) | 1.2 (1.2–1.2) | 2.2 (2.2–2.2) | 73.7 (57.2–82.7) | 0.1 (0.1–0.1) | 0.5 (0.5–0.5) | 0.9 (0.9–1.0) | 1.2 (1.2–1.3) | 55.5 (37.7–74.7) | 1.78 |
| Acute and transient psychotic disorders (F23) | 0.2 (0.2–0.2) | 0.8 (0.7–0.8) | 1.1 (1.1–1.1) | 1.2 (1.2–1.3) | 42.2 (28.4–61.7) | 0.3 (0.3–0.3) | 0.9 (0.8–0.9) | 1.1 (1.0–1.1) | 1.1 (1.1–1.1) | 33.5 (24.3–48.9) | 1.10 |
| Induced delusional disorder (F24) | 0.0 (0.0–0.0) | 0.0 (0.0–0.0) | 0.0 (0.0–0.0) | 0.0 (0.0–0.0) | 51.0 (32.7–71.9) | 0.0 (0.0–0.0) | 0.0 (0.0–0.0) | 0.0 (0.0–0.0) | 0.0 (0.0–0.0) | 41.2 (28.2–65.8) | 1.36 |
| Schizoaffective disorders (F25) | 0.1 (0.1–0.1) | 0.6 (0.5–0.6) | 0.8 (0.8–0.9) | 0.9 (0.9–0.9) | 43.6 (30.3–57.2) | 0.1 (0.1–0.1) | 0.5 (0.4–0.5) | 0.6 (0.6–0.6) | 0.7 (0.6–0.7) | 39.7 (29.0–53.1) | 1.35 |
| Other nonorganic psychotic disorders (F28) | 0.1 (0.1–0.1) | 0.2 (0.1–0.2) | 0.2 (0.2–0.2) | 0.2 (0.2–0.2) | 37.9 (23.7–60.3) | 0.1 (0.1–0.1) | 0.1 (0.1–0.2) | 0.2 (0.2–0.2) | 0.2 (0.2–0.2) | 30.1 (21.4–44.4) | 1.27 |
| Unspecified nonorganic psychosis (F29) | 0.9 (0.9–0.9) | 2.1 (2.1–2.1) | 2.8 (2.8–2.9) | 3.1 (3.1–3.2) | 37.0 (23.2–57.8) | 1.1 (1.1–1.1) | 2.6 (2.6–2.6) | 3.1 (3.0–3.1) | 3.2 (3.2–3.2) | 30.1 (22.3–44.4) | 0.99 |
| **Mood disorders (F30–F39)** | 14.1 (14.1–14.2) | 28.5 (28.4–28.6) | 35.7 (35.6–35.8) | 39.0 (38.9–39.1) | 32.4 (20.4–51.7) | 7.5 (7.4–7.5) | 17.8 (17.8–17.9) | 23.1 (23.1–23.2) | 24.8 (24.7–24.9) | 35.4 (22.9–52.2) | 1.57 |
| Mania and bipolar disorder (F30–F31) | 1.0 (1.0–1.1) | 3.0 (3.0–3.0) | 3.8 (3.8–3.9) | 4.0 (4.0–4.1) | 35.3 (24.8–50.3) | 0.5 (0.5–0.5) | 2.1 (2.0–2.1) | 2.8 (2.8–2.8) | 2.9 (2.9–2.9) | 39.5 (28.2–52.0) | 1.39 |
| Depressive disorders (F32–F33) | 13.5 (13.4–13.5) | 27.2 (27.1–27.3) | 34.3 (34.2–34.4) | 37.4 (37.3–37.5) | 32.7 (20.5–52.0) | 6.9 (6.9–7.0) | 16.6 (16.5–16.6) | 21.6 (21.5–21.7) | 23.2 (23.1–23.2) | 35.6 (22.9–52.5) | 1.62 |
| Others (F34–F39) | 1.7 (1.7–1.8) | 4.0 (3.9–4.0) | 5.4 (5.4–5.5) | 5.9 (5.9–6.0) | 36.7 (23.2–55.4) | 0.9 (0.9–0.9) | 2.5 (2.5–2.5) | 3.3 (3.3–3.4) | 3.5 (3.5–3.5) | 36.7 (24.8–52.6) | 1.69 |
| **Neurotic, stress–related and somatoform disorders (F40–F48)** | 18.7 (18.6–18.7) | 37.5 (37.4–37.6) | 44.9 (44.8–45.0) | 46.6 (46.5–46.7) | 29.3 (19.7–45.1) | 10.5 (10.4–10.5) | 21.8 (21.7–21.9) | 26.1 (26.0–26.2) | 27.0 (26.9–27.1) | 29.7 (20.3–45.1) | 1.72 |
| Anxiety disorders (F40–F41) | 12.7 (12.6–12.7) | 23.7 (23.6–23.8) | 27.8 (27.8–27.9) | 29.1 (29.1–29.2) | 27.6 (19.2–43.5) | 6.7 (6.6–6.7) | 13.9 (13.9–14.0) | 16.3 (16.2–16.4) | 16.8 (16.7–16.9) | 28.7 (20.7–42.8) | 1.74 |
| Obsessive–compulsive disorder (F42) | 1.1 (1.1–1.1) | 1.9 (1.9–1.9) | 2.0 (2.0–2.1) | 2.1 (2.1–2.1) | 24.6 (17.9–34.2) | 0.7 (0.7–0.8) | 1.3 (1.3–1.3) | 1.4 (1.4–1.4) | 1.4 (1.4–1.5) | 24.4 (16.8–35.5) | 1.47 |
| Reaction to severe stress, and adjustment disorders (F43) | 5.3 (5.3–5.4) | 14.8 (14.8–14.9) | 19.2 (19.1–19.3) | 19.9 (19.8–20.0) | 36.1 (24.1–50.3) | 3.1 (3.1–3.2) | 7.4 (7.3–7.4) | 9.2 (9.2–9.3) | 9.6 (9.5–9.6) | 33.8 (21.3–48.6) | 2.08 |
| Dissociative disorders (F44) | 0.3 (0.3–0.4) | 0.7 (0.7–0.7) | 0.8 (0.8–0.8) | 0.8 (0.8–0.8) | 27.9 (19.4–41.9) | 0.1 (0.1–0.1) | 0.2 (0.2–0.2) | 0.2 (0.2–0.2) | 0.2 (0.2–0.3) | 30.1 (19.7–45.7) | 3.32 |
| Somatoform disorders (F45) | 2.2 (2.1–2.2) | 7.5 (7.5–7.6) | 10.1 (10.1–10.2) | 10.6 (10.5–10.7) | 38.2 (26.8–52.5) | 0.8 (0.8–0.9) | 3.0 (3.0–3.0) | 4.1 (4.1–4.2) | 4.4 (4.3–4.4) | 39.7 (27.5–54.0) | 2.43 |
| Other neurotic disorders (F48) | 0.1 (0.1–0.1) | 0.2 (0.2–0.2) | 0.3 (0.3–0.3) | 0.4 (0.4–0.4) | 44.9 (28.3–68.1) | 0.1 (0.0–0.1) | 0.1 (0.1–0.1) | 0.2 (0.2–0.2) | 0.2 (0.2–0.2) | 45.3 (26.3–68.7) | 1.79 |
| **Behavioral syndromes (F50–F59)** | 7.6 (7.5–7.7) | 15.6 (15.5–15.6) | 22.0 (21.9–22.1) | 24.7 (24.7–24.8) | 38.5 (21.9–61.6) | 3.8 (3.7–3.8) | 9.8 (9.7–9.8) | 14.7 (14.6–14.8) | 16.7 (16.6–16.8) | 43.5 (26.4–63.7) | 1.48 |
| Eating disorders (F50) | 2.9 (2.8–2.9) | 3.6 (3.5–3.6) | 3.7 (3.7–3.7) | 3.8 (3.7–3.8) | 18.1 (15.3–24.4) | 0.3 (0.3–0.3) | 0.4 (0.4–0.4) | 0.4 (0.4–0.4) | 0.5 (0.4–0.5) | 19.2 (14.0–41.6) | 8.13 |
| Nonorganic sleep disorders (F51) | 2.6 (2.6–2.6) | 8.4 (8.4–8.5) | 13.5 (13.4–13.6) | 15.8 (15.7–15.9) | 47.5 (30.5–66.8) | 1.9 (1.9–1.9) | 5.9 (5.8–5.9) | 8.9 (8.9–9.0) | 10.4 (10.3–10.4) | 45.1 (28.8–65.2) | 1.52 |
| Disorders associated with the puerperium (F53) | 0.1 (0.1–0.1) | 0.3 (0.3–0.3) | 0.3 (0.3–0.3) | 0.3 (0.3–0.3) | 30.1 (26.0–34.2) | 0.0 (0.0–0.0) | 0.0 (0.0–0.0) | 0.0 (0.0–0.0) | 0.0 (0.0–0.0) | 37.8 (17.0–64.8) | 41.57 |
| Other (F54-F59) | 0.2 (0.2–0.3) | 0.4 (0.4–0.4) | 0.5 (0.5–0.5) | 0.5 (0.5–0.6) | 28.2 (15.8–48.9) | 0.3 (0.3–0.3) | 1.1 (1.1–1.2) | 1.8 (1.7–1.8) | 1.8 (1.8–1.9) | 43.6 (29.2–58.6) | 0.29 |
| **Personality disorders (F60–F69)** | 2.2 (2.2–2.2) | 5.3 (5.2–5.3) | 6.2 (6.2–6.3) | 6.3 (6.3–6.4) | 30.1 (22.5–43.9) | 1.3 (1.3–1.3) | 3.7 (3.6–3.7) | 4.3 (4.3–4.4) | 4.4 (4.4–4.4) | 32.4 (23.7–44.9) | 1.44 |
| Paranoid personality disorder (F60.0) | 0.0 (0.0–0.1) | 0.2 (0.2–0.2) | 0.2 (0.2–0.2) | 0.2 (0.2–0.3) | 39.4 (27.3–52.7) | 0.0 (0.0–0.0) | 0.2 (0.1–0.2) | 0.2 (0.2–0.2) | 0.2 (0.2–0.2) | 40.0 (29.6–51.0) | 1.14 |
| Schizoid personality disorder (F60.1) | 0.0 (0.0–0.0) | 0.1 (0.1–0.1) | 0.1 (0.1–0.1) | 0.1 (0.1–0.1) | 37.7 (26.3–49.9) | 0.1 (0.0–0.1) | 0.2 (0.2–0.2) | 0.2 (0.2–0.2) | 0.2 (0.2–0.2) | 33.9 (25.4–46.3) | 0.55 |
| Dissocial personality disorder (F60.2) | 0.0 (0.0–0.0) | 0.1 (0.1–0.1) | 0.1 (0.1–0.1) | 0.1 (0.1–0.1) | 31.0 (25.7–40.7) | 0.1 (0.1–0.1) | 0.3 (0.3–0.3) | 0.4 (0.3–0.4) | 0.4 (0.3–0.4) | 31.9 (25.1–39.7) | 0.23 |
| Emotionally unstable personality disorder (F60.3) | 1.1 (1.1–1.1) | 2.6 (2.6–2.6) | 2.8 (2.8–2.9) | 2.8 (2.8–2.9) | 27.6 (22.1–37.3) | 0.2 (0.2–0.2) | 0.9 (0.9–0.9) | 1.0 (1.0–1.0) | 1.0 (1.0–1.0) | 32.7 (25.3–42.0) | 2.89 |
| Histrionic personality disorder (F60.4) | 0.0 (0.0–0.0) | 0.0 (0.0–0.0) | 0.0 (0.0–0.0) | 0.0 (0.0–0.1) | 41.1 (27.2–53.8) | 0.0 (0.0–0.0) | 0.0 (0.0–0.0) | 0.0 (0.0–0.0) | 0.0 (0.0–0.0) | 39.2 (26.9–51.4) | 2.71 |
| Anankastic personality disorder (F60.5) | 0.1 (0.1–0.1) | 0.6 (0.5–0.6) | 0.7 (0.7–0.8) | 0.7 (0.7–0.8) | 39.0 (29.0–50.7) | 0.0 (0.0–0.0) | 0.2 (0.2–0.3) | 0.4 (0.4–0.4) | 0.4 (0.4–0.4) | 44.2 (33.7–52.9) | 2.03 |
| Anxious [avoidant] personality disorder (F60.6) | 0.1 (0.1–0.1) | 0.3 (0.3–0.3) | 0.4 (0.4–0.4) | 0.4 (0.4–0.4) | 31.0 (23.9–44.3) | 0.1 (0.1–0.1) | 0.3 (0.3–0.3) | 0.4 (0.3–0.4) | 0.4 (0.3–0.4) | 31.1 (24.6–42.7) | 1.04 |
| Dependent personality disorder (F60.7) | 0.0 (0.0–0.0) | 0.2 (0.2–0.2) | 0.3 (0.3–0.3) | 0.3 (0.3–0.3) | 44.7 (32.0–53.8) | 0.0 (0.0–0.0) | 0.1 (0.1–0.1) | 0.1 (0.1–0.1) | 0.1 (0.1–0.1) | 41.3 (31.8–50.8) | 3.05 |
| Other specific personality disorders (F60.8) | 0.1 (0.1–0.1) | 0.4 (0.4–0.4) | 0.5 (0.4–0.5) | 0.5 (0.4–0.5) | 32.7 (24.2–46.0) | 0.1 (0.1–0.1) | 0.3 (0.3–0.3) | 0.4 (0.4–0.4) | 0.4 (0.4–0.4) | 34.1 (24.8–46.2) | 1.12 |
| Personality disorder, unspecified (F60.9) | 0.4 (0.3–0.4) | 1.0 (1.0–1.0) | 1.2 (1.1–1.2) | 1.2 (1.2–1.2) | 31.9 (23.6–44.7) | 0.2 (0.2–0.2) | 0.7 (0.7–0.7) | 0.8 (0.8–0.9) | 0.9 (0.8–0.9) | 34.5 (25.5–45.3) | 1.36 |
| Mixed and other personality disorders (F61) | 0.3 (0.3–0.3) | 1.2 (1.1–1.2) | 1.5 (1.5–1.5) | 1.5 (1.5–1.5) | 36.2 (26.7–48.3) | 0.2 (0.2–0.2) | 1.0 (1.0–1.0) | 1.2 (1.2–1.2) | 1.2 (1.2–1.3) | 37.1 (27.7–47.2) | 1.22 |
| Enduring personality changes, not attributable to brain damage and disease (F62) | 0.0 (0.0–0.0) | 0.0 (0.0–0.0) | 0.1 (0.1–0.1) | 0.1 (0.1–0.1) | 43.4 (28.5–55.4) | 0.0 (0.0–0.0) | 0.0 (0.0–0.0) | 0.1 (0.1–0.1) | 0.1 (0.1–0.1) | 44.7 (27.0–55.8) | 1.03 |
| Others (F63–F69) | 0.4 (0.4–0.4) | 0.7 (0.7–0.7) | 0.8 (0.7–0.8) | 0.8 (0.8–0.8) | 23.7 (17.8–36.3) | 0.4 (0.4–0.5) | 0.8 (0.8–0.8) | 0.9 (0.9–0.9) | 0.9 (0.9–0.9) | 25.4 (19.1–37.2) | 0.87 |
| **Intellectual disability (F70–F79)** | 1.0 (0.9–1.0) | 1.3 (1.2–1.3) | 1.5 (1.5–1.5) | 1.6 (1.6–1.6) | 17.0 (7.0–46.1) | 1.4 (1.4–1.5) | 1.8 (1.7–1.8) | 2.1 (2.0–2.1) | 2.1 (2.1–2.2) | 13.6 (6.0–35.8) | 0.75 |
| Mild intellectual disability (F70) | 0.5 (0.5–0.6) | 0.7 (0.7–0.7) | 0.9 (0.8–0.9) | 0.9 (0.9–0.9) | 17.1 (9.4–40.8) | 0.8 (0.8–0.8) | 1.0 (1.0–1.0) | 1.1 (1.1–1.1) | 1.1 (1.1–1.2) | 15.1 (7.5–32.4) | 0.78 |
| Moderate intellectual disability (F71) | 0.2 (0.2–0.2) | 0.3 (0.2–0.3) | 0.3 (0.3–0.3) | 0.3 (0.3–0.4) | 23.7 (13.6–49.9) | 0.3 (0.2–0.3) | 0.4 (0.3–0.4) | 0.4 (0.4–0.5) | 0.5 (0.4–0.5) | 21.1 (11.6–46.6) | 0.76 |
| Severe intellectual disability (F72) | 0.1 (0.1–0.1) | 0.1 (0.1–0.1) | 0.2 (0.2–0.2) | 0.2 (0.2–0.2) | 30.0 (14.5–55.5) | 0.1 (0.1–0.1) | 0.2 (0.2–0.2) | 0.2 (0.2–0.2) | 0.2 (0.2–0.2) | 26.9 (14.1–51.3) | 0.77 |
| Profound intellectual disability (F73) | 0.0 (0.0–0.0) | 0.1 (0.1–0.1) | 0.1 (0.1–0.1) | 0.1 (0.1–0.1) | 32.7 (14.9–50.2) | 0.0 (0.0–0.0) | 0.1 (0.1–0.1) | 0.1 (0.1–0.1) | 0.1 (0.1–0.1) | 33.3 (14.6–51.0) | 0.89 |
| Other intellectual disability (F78) | 0.0 (0.0–0.0) | 0.0 (0.0–0.0) | 0.1 (0.0–0.1) | 0.1 (0.1–0.1) | 36.2 (15.0–57.1) | 0.0 (0.0–0.0) | 0.0 (0.0–0.0) | 0.1 (0.1–0.1) | 0.1 (0.1–0.1) | 32.9 (12.0–56.9) | 0.84 |
| Unspecified intellectual disability (F79) | 0.4 (0.4–0.4) | 0.5 (0.5–0.5) | 0.6 (0.5–0.6) | 0.6 (0.6–0.6) | 10.5 (4.7–33.6) | 0.7 (0.6–0.7) | 0.8 (0.7–0.8) | 0.8 (0.8–0.9) | 0.9 (0.8–0.9) | 7.9 (4.8–20.3) | 0.67 |
| **Disorders of psychological development (F80–F89)** | 7.4 (7.3–7.5) | 7.8 (7.8–7.9) | 8.0 (7.9–8.1) | 8.1 (8.1–8.2) | 6.9 (5.0–13.3) | 14.2 (14.1–14.2) | 14.6 (14.5–14.7) | 14.7 (14.7–14.8) | 14.8 (14.7–14.9) | 6.2 (4.5–9.9) | 0.55 |
| Specific developmental disorders of speech and language (F80) | 2.6 (2.6–2.7) | 2.7 (2.6–2.7) | 2.8 (2.7–2.8) | 2.9 (2.9–3.0) | 5.6 (4.3–7.8) | 5.8 (5.7–5.8) | 5.8 (5.8–5.9) | 5.9 (5.9–6.0) | 6.0 (5.9–6.0) | 5.2 (3.9–6.7) | 0.49 |
| Specific developmental disorders of scholastic skills (F81) | 1.9 (1.8–1.9) | 2.0 (2.0–2.1) | 2.1 (2.0–2.1) | 2.1 (2.0–2.1) | 12.3 (9.2–16.5) | 3.3 (3.2–3.3) | 3.4 (3.4–3.5) | 3.5 (3.4–3.5) | 3.5 (3.4–3.5) | 10.9 (8.7–14.5) | 0.59 |
| Specific developmental disorder of motor function (F82) | 0.7 (0.7–0.7) | 0.7 (0.7–0.7) | 0.7 (0.7–0.7) | 0.7 (0.7–0.7) | 5.4 (3.4–7.1) | 1.8 (1.8–1.8) | 1.8 (1.8–1.8) | 1.8 (1.8–1.8) | 1.8 (1.8–1.8) | 5.8 (4.7–7.3) | 0.38 |
| Mixed specific developmental disorders (F83) | 1.3 (1.3–1.4) | 1.4 (1.3–1.4) | 1.4 (1.3–1.4) | 1.4 (1.3–1.4) | 6.1 (4.5–9.2) | 2.9 (2.9–2.9) | 2.9 (2.9–3.0) | 2.9 (2.9–3.0) | 2.9 (2.9–3.0) | 5.8 (4.4–8.1) | 0.46 |
| Pervasive developmental disorders (F84) | 0.8 (0.8–0.8) | 1.0 (1.0–1.0) | 1.0 (1.0–1.0) | 1.0 (1.0–1.0) | 13.3 (7.7–20.3) | 2.3 (2.3–2.3) | 2.5 (2.5–2.6) | 2.6 (2.5–2.6) | 2.6 (2.5–2.6) | 9.9 (6.1–15.2) | 0.39 |
| Other disorders of psychological development (F88) | 0.2 (0.2–0.2) | 0.2 (0.2–0.3) | 0.2 (0.2–0.3) | 0.2 (0.2–0.3) | 8.7 (6.2–11.8) | 0.7 (0.6–0.7) | 0.7 (0.6–0.7) | 0.7 (0.6–0.7) | 0.7 (0.6–0.7) | 8.2 (6.2–10.7) | 0.37 |
| Unspecified disorder of psychological development (F89) | 0.1 (0.1–0.1) | 0.1 (0.1–0.2) | 0.2 (0.1–0.2) | 0.2 (0.1–0.2) | 10.2 (6.4–15.8) | 0.3 (0.3–0.3) | 0.3 (0.3–0.4) | 0.4 (0.3–0.4) | 0.4 (0.3–0.4) | 8.6 (6.2–11.8) | 0.43 |
| **Behavioral and emotional disorders (F90–F98)** | 12.9 (12.8–13.0) | 14.9 (14.8–15.0) | 15.5 (15.4–15.6) | 15.8 (15.7–15.8) | 12.3 (6.3–18.0) | 19.1 (19.0–19.2) | 20.6 (20.5–20.7) | 20.9 (20.9–21.0) | 21.1 (21.0–21.2) | 8.1 (5.5–12.9) | 0.75 |
| ADHD (F90) | 2.4 (2.4–2.5) | 3.5 (3.4–3.5) | 3.5 (3.5–3.6) | 3.5 (3.5–3.6) | 17.2 (9.7–28.3) | 6.8 (6.7–6.8) | 7.8 (7.7–7.8) | 7.8 (7.8–7.9) | 7.9 (7.8–7.9) | 10.0 (7.6–15.3) | 0.45 |
| Conduct disorders (F91) | 0.8 (0.7–0.8) | 0.8 (0.7–0.8) | 0.8 (0.8–0.8) | 0.8 (0.8–0.8) | 14.0 (9.8–15.7) | 1.7 (1.7–1.7) | 1.7 (1.7–1.8) | 1.8 (1.7–1.8) | 1.8 (1.8–1.8) | 11.6 (8.2–14.8) | 0.45 |
| Mixed disorders of conduct and emotions (F92) | 1.2 (1.1–1.2) | 1.2 (1.1–1.2) | 1.2 (1.1–1.2) | 1.2 (1.2–1.2) | 13.8 (10.3–15.5) | 2.3 (2.3–2.4) | 2.3 (2.3–2.4) | 2.3 (2.3–2.4) | 2.3 (2.3–2.4) | 10.6 (8.4–13.1) | 0.50 |
| Emotional disorders with onset specific to childhood (F93) | 3.1 (3.1–3.2) | 3.2 (3.1–3.2) | 3.2 (3.2–3.2) | 3.2 (3.2–3.3) | 14.0 (10.6–16.3) | 2.7 (2.7–2.8) | 2.8 (2.8–2.8) | 2.8 (2.8–2.8) | 2.8 (2.8–2.9) | 10.8 (8.4–13.8) | 1.15 |
| Disorders of social functioning with onset specific to childhood and adolescence (F94) | 0.5 (0.5–0.6) | 0.6 (0.5–0.6) | 0.6 (0.5–0.6) | 0.6 (0.5–0.6) | 8.7 (6.1–12.2) | 0.7 (0.7–0.7) | 0.7 (0.7–0.7) | 0.7 (0.7–0.7) | 0.7 (0.7–0.7) | 8.5 (6.0–11.5) | 0.80 |
| Tic disorders (F95) | 0.4 (0.4–0.4) | 0.4 (0.4–0.4) | 0.4 (0.4–0.4) | 0.4 (0.4–0.5) | 10.0 (7.0–15.6) | 1.1 (1.0–1.1) | 1.1 (1.1–1.1) | 1.1 (1.1–1.2) | 1.1 (1.1–1.2) | 9.1 (6.9–11.9) | 0.38 |
| Other behavioral and emotional disorders with onset usually occurring in childhood and adolescence (F98) | 3.2 (3.2–3.2) | 3.3 (3.3–3.4) | 3.4 (3.3–3.4) | 3.4 (3.4–3.5) | 7.5 (5.5–13.0) | 4.7 (4.7–4.8) | 4.8 (4.8–4.9) | 4.9 (4.8–4.9) | 4.9 (4.8–4.9) | 7.3 (5.6–10.4) | 0.70 |
| *^1^Women to men ratio of the cumulative incidence estimates at the age of 100.* | | | | | | | | | | | |

### Supplementary Fig. 2: Cumulative incidence of mental disorders by gender and ICD–10 sub-chapter category

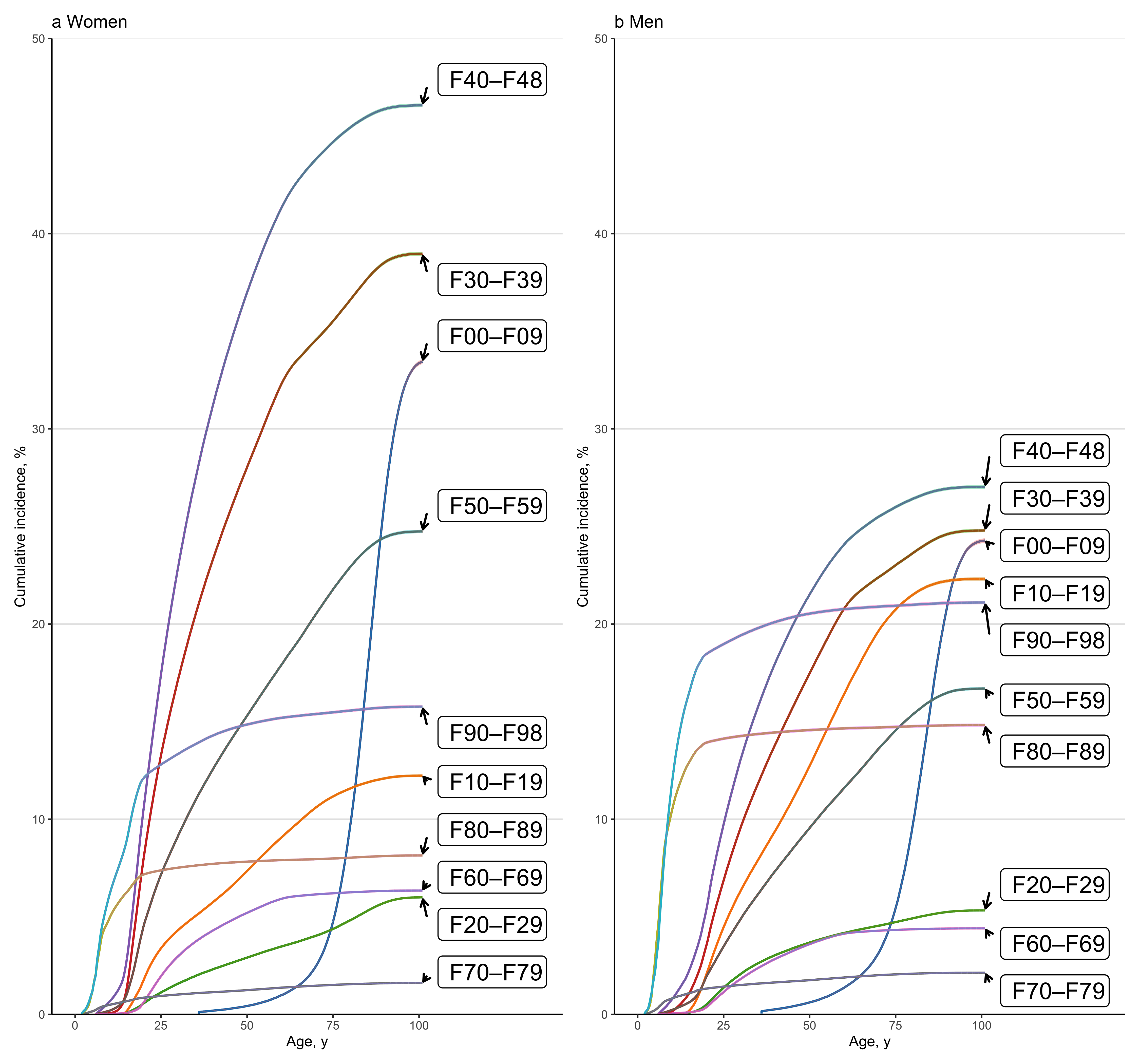

### Supplementary Table 5: 12-month service utilization for medical contacts with diagnosed mental disorders by gender, age group, and diagnosis in 2019^1^

|  | **Number of individuals and service utilization by agegroup, N (%)** | | | | | | | | | |
| --- | --- | --- | --- | --- | --- | --- | --- | --- | --- | --- |
|  | **Women** | | | | | **Men** | | | | |
|  | **0–24** | **25–49** | **50–74** | **75–99** | **0–99** | **0–24** | **25–49** | **50–74** | **75–99** | **0–99** |
| **Any mental disorder** | 71 436 (9.9) | 86 927 (10.4) | 62 322 (6.8) | 31 833 (10.0) | 252 518 (9.0) | 78 841 (10.4) | 63 607 (7.2) | 50 760 (5.8) | 17 739 (8.6) | 210 947 (7.7) |
| Non-organic mental disorders (F10–F99) | 71 411 (9.9) | 86 835 (10.3) | 59 479 (6.5) | 15 026 (4.7) | 232 751 (8.3) | 78 823 (10.4) | 63 466 (7.1) | 47 682 (5.5) | 7 247 (3.5) | 197 218 (7.2) |
| **Organic mental disorders (F00–F09)** | 0 (0.0) | 193 (0.0) | 3 866 (0.4) | 18 757 (5.9) | 22 816 (0.8) | 0 (0.0) | 286 (0.0) | 4 237 (0.5) | 11 414 (5.5) | 15 937 (0.6) |
| Dementias (F00–03) | 0 (0.0) | 17 (0.0) | 2 782 (0.3) | 16 212 (5.1) | 19 011 (0.7) | 0 (0.0) | 15 (0.0) | 2 711 (0.3) | 9 672 (4.7) | 12 398 (0.5) |
| Others (F04–09) | 0 (0.0) | 158 (0.0) | 1 274 (0.1) | 3 373 (1.1) | 4 805 (0.2) | 0 (0.0) | 261 (0.0) | 1 726 (0.2) | 2 249 (1.1) | 4 236 (0.2) |
| **Substance use disorders (F10–F19)** | 3 254 (0.5) | 6 997 (0.8) | 7 603 (0.8) | 969 (0.3) | 18 823 (0.7) | 4 700 (0.6) | 16 473 (1.9) | 17 320 (2.0) | 1 783 (0.9) | 40 276 (1.5) |
| Alcohol use disorders (F10) | 1 741 (0.2) | 3 093 (0.4) | 4 619 (0.5) | 597 (0.2) | 10 050 (0.4) | 2 100 (0.3) | 8 366 (0.9) | 13 056 (1.5) | 1 341 (0.6) | 24 863 (0.9) |
| Opiods use disorders (F11) | 209 (0.0) | 1 366 (0.2) | 124 (0.0) | 8 (0.0) | 1 707 (0.1) | 249 (0.0) | 2 953 (0.3) | 297 (0.0) | 0 (0.0) | 3 499 (0.1) |
| Cannabinoids use disorders (F12) | 272 (0.0) | 225 (0.0) | 4 (0.0) | 0 (0.0) | 501 (0.0) | 732 (0.1) | 1 013 (0.1) | 48 (0.0) | 0 (0.0) | 1 793 (0.1) |
| Sedatives or hypnotics use disorders (F13) | 131 (0.0) | 575 (0.1) | 546 (0.1) | 106 (0.0) | 1 358 (0.0) | 248 (0.0) | 1 412 (0.2) | 531 (0.1) | 47 (0.0) | 2 238 (0.1) |
| Cocaine use disorders (F14) | 0 (0.0) | 0 (0.0) | 0 (0.0) | 0 (0.0) | 0 (0.0) | 4 (0.0) | 0 (0.0) | 0 (0.0) | 0 (0.0) | 4 (0.0) |
| Other stimulants use disorders (F15) | 175 (0.0) | 270 (0.0) | 14 (0.0) | 0 (0.0) | 459 (0.0) | 238 (0.0) | 805 (0.1) | 38 (0.0) | 0 (0.0) | 1 081 (0.0) |
| Hallucinogens use disorders (F16) | 11 (0.0) | 0 (0.0) | 0 (0.0) | 0 (0.0) | 11 (0.0) | 61 (0.0) | 39 (0.0) | 0 (0.0) | 0 (0.0) | 100 (0.0) |
| Tobacco use disorders (F17) | 110 (0.0) | 760 (0.1) | 1 349 (0.1) | 119 (0.0) | 2 338 (0.1) | 98 (0.0) | 807 (0.1) | 1 669 (0.2) | 153 (0.1) | 2 727 (0.1) |
| Volatile solvents use disorders (F18) | 0 (0.0) | 0 (0.0) | 0 (0.0) | 0 (0.0) | 0 (0.0) | 0 (0.0) | 0 (0.0) | 0 (0.0) | 0 (0.0) | 0 (0.0) |
| Multiple and other substances use disorders (F19) | 608 (0.1) | 877 (0.1) | 123 (0.0) | 0 (0.0) | 1 608 (0.1) | 907 (0.1) | 2 509 (0.3) | 216 (0.0) | 0 (0.0) | 3 632 (0.1) |
| **Schizophrenia spectrum (F20–F29)** | 1 364 (0.2) | 6 307 (0.8) | 8 204 (0.9) | 2 322 (0.7) | 18 197 (0.7) | 1 665 (0.2) | 9 750 (1.1) | 6 947 (0.8) | 706 (0.3) | 19 068 (0.7) |
| Schizophrenia (F20) | 233 (0.0) | 2 490 (0.3) | 4 031 (0.4) | 811 (0.3) | 7 565 (0.3) | 364 (0.0) | 5 033 (0.6) | 4 195 (0.5) | 320 (0.2) | 9 912 (0.4) |
| Schizotypal disorder (F21) | 18 (0.0) | 179 (0.0) | 115 (0.0) | 4 (0.0) | 316 (0.0) | 10 (0.0) | 335 (0.0) | 118 (0.0) | 0 (0.0) | 463 (0.0) |
| Persistent delusional disorders (F22) | 14 (0.0) | 360 (0.0) | 1 098 (0.1) | 1 015 (0.3) | 2 487 (0.1) | 34 (0.0) | 542 (0.1) | 630 (0.1) | 200 (0.1) | 1 406 (0.1) |
| Acute and transient psychotic disorders (F23) | 105 (0.0) | 450 (0.1) | 258 (0.0) | 63 (0.0) | 876 (0.0) | 152 (0.0) | 466 (0.1) | 138 (0.0) | 9 (0.0) | 765 (0.0) |
| Induced delusional disorder (F24) | 0 (0.0) | 0 (0.0) | 0 (0.0) | 0 (0.0) | 0 (0.0) | 0 (0.0) | 0 (0.0) | 0 (0.0) | 0 (0.0) | 0 (0.0) |
| Schizoaffective disorders (F25) | 142 (0.0) | 1 339 (0.2) | 1 561 (0.2) | 149 (0.0) | 3 191 (0.1) | 64 (0.0) | 1 227 (0.1) | 850 (0.1) | 41 (0.0) | 2 182 (0.1) |
| Other nonorganic psychotic disorders (F28) | 49 (0.0) | 127 (0.0) | 34 (0.0) | 4 (0.0) | 214 (0.0) | 32 (0.0) | 89 (0.0) | 5 (0.0) | 0 (0.0) | 126 (0.0) |
| Unspecified nonorganic psychosis (F29) | 949 (0.1) | 1 996 (0.2) | 1 100 (0.1) | 243 (0.1) | 4 288 (0.2) | 1 200 (0.2) | 2 879 (0.3) | 765 (0.1) | 70 (0.0) | 4 914 (0.2) |
| **Mood disorders (F30–F39)** | 22 491 (3.1) | 39 107 (4.7) | 22 901 (2.5) | 5 400 (1.7) | 89 899 (3.2) | 10 197 (1.3) | 22 338 (2.5) | 13 607 (1.6) | 1 923 (0.9) | 48 065 (1.8) |
| Mania and bipolar disorder (F30–F31) | 1 200 (0.2) | 5 589 (0.7) | 3 559 (0.4) | 406 (0.1) | 10 754 (0.4) | 541 (0.1) | 3 373 (0.4) | 2 816 (0.3) | 245 (0.1) | 6 975 (0.3) |
| Depressive disorders (F32–F33) | 20 733 (2.9) | 32 852 (3.9) | 18 452 (2.0) | 4 635 (1.5) | 76 672 (2.7) | 9 225 (1.2) | 18 261 (2.1) | 10 321 (1.2) | 1 573 (0.8) | 39 380 (1.4) |
| Others (F34–F39) | 1 908 (0.3) | 3 074 (0.4) | 1 978 (0.2) | 513 (0.2) | 7 473 (0.3) | 944 (0.1) | 2 218 (0.2) | 1 137 (0.1) | 156 (0.1) | 4 455 (0.2) |
| **Neurotic, stress–related and somatoform disorders (F40–F48)** | 30 556 (4.2) | 46 384 (5.5) | 20 042 (2.2) | 3 433 (1.1) | 100 415 (3.6) | 14 167 (1.9) | 23 007 (2.6) | 8 773 (1.0) | 972 (0.5) | 46 919 (1.7) |
| Anxiety disorders (F40–F41) | 21 928 (3.0) | 26 430 (3.1) | 9 422 (1.0) | 2 101 (0.7) | 59 881 (2.1) | 9 547 (1.3) | 15 000 (1.7) | 4 909 (0.6) | 534 (0.3) | 29 990 (1.1) |
| Obsessive–compulsive disorder (F42) | 1 899 (0.3) | 2 196 (0.3) | 374 (0.0) | 33 (0.0) | 4 502 (0.2) | 1 359 (0.2) | 1 451 (0.2) | 307 (0.0) | 6 (0.0) | 3 123 (0.1) |
| Reaction to severe stress, and adjustment disorders (F43) | 4 891 (0.7) | 11 043 (1.3) | 5 456 (0.6) | 584 (0.2) | 21 974 (0.8) | 2 443 (0.3) | 3 985 (0.4) | 1 802 (0.2) | 185 (0.1) | 8 415 (0.3) |
| Dissociative disorders (F44) | 501 (0.1) | 916 (0.1) | 165 (0.0) | 0 (0.0) | 1 582 (0.1) | 100 (0.0) | 171 (0.0) | 27 (0.0) | 0 (0.0) | 298 (0.0) |
| Somatoform disorders (F45) | 2 267 (0.3) | 7 566 (0.9) | 4 211 (0.5) | 525 (0.2) | 14 569 (0.5) | 851 (0.1) | 2 824 (0.3) | 1 582 (0.2) | 172 (0.1) | 5 429 (0.2) |
| Other neurotic disorders (F48) | 13 (0.0) | 104 (0.0) | 22 (0.0) | 0 (0.0) | 139 (0.0) | 0 (0.0) | 17 (0.0) | 4 (0.0) | 0 (0.0) | 21 (0.0) |
| **Behavioral syndromes (F50–F59)** | 8 220 (1.1) | 11 619 (1.4) | 11 511 (1.3) | 4 249 (1.3) | 35 599 (1.3) | 4 011 (0.5) | 7 628 (0.9) | 8 250 (0.9) | 2 274 (1.1) | 22 163 (0.8) |
| Eating disorders (F50) | 3 520 (0.5) | 1 848 (0.2) | 204 (0.0) | 4 (0.0) | 5 576 (0.2) | 328 (0.0) | 115 (0.0) | 12 (0.0) | 0 (0.0) | 455 (0.0) |
| Nonorganic sleep disorders (F51) | 2 397 (0.3) | 6 423 (0.8) | 7 017 (0.8) | 3 015 (1.0) | 18 852 (0.7) | 1 852 (0.2) | 4 152 (0.5) | 3 807 (0.4) | 1 303 (0.6) | 11 114 (0.4) |
| Disorders associated with the puerperium (F53) | 18 (0.0) | 130 (0.0) | 0 (0.0) | 0 (0.0) | 148 (0.0) | 0 (0.0) | 0 (0.0) | 0 (0.0) | 0 (0.0) | 0 (0.0) |
| Other (F54-F59) | 84 (0.0) | 17 (0.0) | 8 (0.0) | 0 (0.0) | 109 (0.0) | 77 (0.0) | 8 (0.0) | 0 (0.0) | 0 (0.0) | 85 (0.0) |
| **Personality disorders (F60–F69)** | 3 334 (0.5) | 7 937 (0.9) | 2 172 (0.2) | 105 (0.0) | 13 548 (0.5) | 1 203 (0.2) | 4 364 (0.5) | 1 360 (0.2) | 47 (0.0) | 6 974 (0.3) |
| Paranoid personality disorder (F60.0) | 20 (0.0) | 51 (0.0) | 16 (0.0) | 0 (0.0) | 87 (0.0) | 4 (0.0) | 65 (0.0) | 20 (0.0) | 0 (0.0) | 89 (0.0) |
| Schizoid personality disorder (F60.1) | 0 (0.0) | 9 (0.0) | 0 (0.0) | 0 (0.0) | 9 (0.0) | 10 (0.0) | 152 (0.0) | 17 (0.0) | 0 (0.0) | 179 (0.0) |
| Dissocial personality disorder (F60.2) | 7 (0.0) | 12 (0.0) | 0 (0.0) | 0 (0.0) | 19 (0.0) | 45 (0.0) | 316 (0.0) | 50 (0.0) | 0 (0.0) | 411 (0.0) |
| Emotionally unstable personality disorder (F60.3) | 1 573 (0.2) | 4 246 (0.5) | 738 (0.1) | 0 (0.0) | 6 557 (0.2) | 150 (0.0) | 922 (0.1) | 219 (0.0) | 0 (0.0) | 1 291 (0.0) |
| Histrionic personality disorder (F60.4) | 0 (0.0) | 0 (0.0) | 0 (0.0) | 0 (0.0) | 0 (0.0) | 0 (0.0) | 0 (0.0) | 0 (0.0) | 0 (0.0) | 0 (0.0) |
| Anankastic personality disorder (F60.5) | 114 (0.0) | 747 (0.1) | 274 (0.0) | 0 (0.0) | 1 135 (0.0) | 21 (0.0) | 282 (0.0) | 138 (0.0) | 0 (0.0) | 441 (0.0) |
| Anxious [avoidant] personality disorder (F60.6) | 154 (0.0) | 418 (0.0) | 38 (0.0) | 0 (0.0) | 610 (0.0) | 92 (0.0) | 416 (0.0) | 65 (0.0) | 0 (0.0) | 573 (0.0) |
| Dependent personality disorder (F60.7) | 17 (0.0) | 75 (0.0) | 68 (0.0) | 0 (0.0) | 160 (0.0) | 0 (0.0) | 0 (0.0) | 0 (0.0) | 0 (0.0) | 0 (0.0) |
| Other specific personality disorders (F60.8) | 97 (0.0) | 347 (0.0) | 99 (0.0) | 0 (0.0) | 543 (0.0) | 54 (0.0) | 256 (0.0) | 62 (0.0) | 0 (0.0) | 372 (0.0) |
| Personality disorder, unspecified (F60.9) | 244 (0.0) | 556 (0.1) | 181 (0.0) | 4 (0.0) | 985 (0.0) | 91 (0.0) | 417 (0.0) | 120 (0.0) | 0 (0.0) | 628 (0.0) |
| Mixed and other personality disorders (F61) | 242 (0.0) | 1 204 (0.1) | 485 (0.1) | 18 (0.0) | 1 949 (0.1) | 76 (0.0) | 911 (0.1) | 377 (0.0) | 0 (0.0) | 1 364 (0.0) |
| Enduring personality changes, not attributable to brain damage and disease (F62) | 0 (0.0) | 0 (0.0) | 4 (0.0) | 0 (0.0) | 4 (0.0) | 0 (0.0) | 13 (0.0) | 0 (0.0) | 0 (0.0) | 13 (0.0) |
| Others (F63–F69) | 1 046 (0.1) | 767 (0.1) | 116 (0.0) | 0 (0.0) | 1 929 (0.1) | 666 (0.1) | 962 (0.1) | 148 (0.0) | 0 (0.0) | 1 776 (0.1) |
| **Intellectual disability (F70–F79)** | 2 604 (0.4) | 1 339 (0.2) | 1 027 (0.1) | 108 (0.0) | 5 078 (0.2) | 4 128 (0.5) | 1 600 (0.2) | 1 159 (0.1) | 103 (0.0) | 6 990 (0.3) |
| Mild intellectual disability (F70) | 1 079 (0.1) | 564 (0.1) | 428 (0.0) | 28 (0.0) | 2 099 (0.1) | 1 578 (0.2) | 612 (0.1) | 437 (0.0) | 27 (0.0) | 2 654 (0.1) |
| Moderate intellectual disability (F71) | 345 (0.0) | 262 (0.0) | 212 (0.0) | 10 (0.0) | 829 (0.0) | 493 (0.1) | 354 (0.0) | 253 (0.0) | 4 (0.0) | 1 104 (0.0) |
| Severe intellectual disability (F72) | 160 (0.0) | 139 (0.0) | 79 (0.0) | 0 (0.0) | 378 (0.0) | 273 (0.0) | 193 (0.0) | 117 (0.0) | 0 (0.0) | 583 (0.0) |
| Profound intellectual disability (F73) | 44 (0.0) | 67 (0.0) | 41 (0.0) | 0 (0.0) | 152 (0.0) | 65 (0.0) | 58 (0.0) | 44 (0.0) | 0 (0.0) | 167 (0.0) |
| Other intellectual disability (F78) | 0 (0.0) | 0 (0.0) | 0 (0.0) | 0 (0.0) | 0 (0.0) | 0 (0.0) | 4 (0.0) | 0 (0.0) | 0 (0.0) | 4 (0.0) |
| Unspecified intellectual disability (F79) | 967 (0.1) | 196 (0.0) | 120 (0.0) | 4 (0.0) | 1 287 (0.0) | 1 685 (0.2) | 262 (0.0) | 166 (0.0) | 10 (0.0) | 2 123 (0.1) |
| **Disorders of psychological development (F80–F89)** | 12 798 (1.8) | 1 445 (0.2) | 286 (0.0) | 143 (0.0) | 14 672 (0.5) | 27 600 (3.6) | 2 227 (0.3) | 403 (0.0) | 63 (0.0) | 30 293 (1.1) |
| Specific developmental disorders of speech and language (F80) | 3 164 (0.4) | 129 (0.0) | 103 (0.0) | 139 (0.0) | 3 535 (0.1) | 8 095 (1.1) | 184 (0.0) | 110 (0.0) | 60 (0.0) | 8 449 (0.3) |
| Specific developmental disorders of scholastic skills (F81) | 2 805 (0.4) | 430 (0.1) | 53 (0.0) | 0 (0.0) | 3 288 (0.1) | 4 709 (0.6) | 432 (0.0) | 91 (0.0) | 0 (0.0) | 5 232 (0.2) |
| Specific developmental disorder of motor function (F82) | 695 (0.1) | 4 (0.0) | 0 (0.0) | 0 (0.0) | 699 (0.0) | 2 197 (0.3) | 9 (0.0) | 0 (0.0) | 0 (0.0) | 2 206 (0.1) |
| Mixed specific developmental disorders (F83) | 2 073 (0.3) | 85 (0.0) | 0 (0.0) | 0 (0.0) | 2 158 (0.1) | 4 691 (0.6) | 156 (0.0) | 0 (0.0) | 0 (0.0) | 4 847 (0.2) |
| Pervasive developmental disorders (F84) | 2 130 (0.3) | 705 (0.1) | 54 (0.0) | 0 (0.0) | 2 889 (0.1) | 6 145 (0.8) | 1 412 (0.2) | 122 (0.0) | 0 (0.0) | 7 679 (0.3) |
| Other disorders of psychological development (F88) | 514 (0.1) | 0 (0.0) | 0 (0.0) | 0 (0.0) | 514 (0.0) | 1 561 (0.2) | 0 (0.0) | 0 (0.0) | 0 (0.0) | 1 561 (0.1) |
| Unspecified disorder of psychological development (F89) | 176 (0.0) | 0 (0.0) | 0 (0.0) | 0 (0.0) | 176 (0.0) | 590 (0.1) | 4 (0.0) | 0 (0.0) | 0 (0.0) | 594 (0.0) |
| **Behavioral and emotional disorders (F90–F98)** | 21 102 (2.9) | 5 068 (0.6) | 823 (0.1) | 159 (0.1) | 27 152 (1.0) | 39 637 (5.2) | 4 644 (0.5) | 736 (0.1) | 70 (0.0) | 45 087 (1.7) |
| ADHD (F90) | 6 555 (0.9) | 3 758 (0.4) | 314 (0.0) | 0 (0.0) | 10 627 (0.4) | 19 536 (2.6) | 3 644 (0.4) | 344 (0.0) | 0 (0.0) | 23 524 (0.9) |
| Conduct disorders (F91) | 670 (0.1) | 0 (0.0) | 0 (0.0) | 4 (0.0) | 674 (0.0) | 1 944 (0.3) | 0 (0.0) | 0 (0.0) | 0 (0.0) | 1 944 (0.1) |
| Mixed disorders of conduct and emotions (F92) | 1 140 (0.2) | 0 (0.0) | 0 (0.0) | 0 (0.0) | 1 140 (0.0) | 2 869 (0.4) | 4 (0.0) | 0 (0.0) | 0 (0.0) | 2 873 (0.1) |
| Emotional disorders with onset specific to childhood (F93) | 4 781 (0.7) | 15 (0.0) | 0 (0.0) | 0 (0.0) | 4 796 (0.2) | 4 504 (0.6) | 0 (0.0) | 0 (0.0) | 0 (0.0) | 4 504 (0.2) |
| Disorders of social functioning with onset specific to childhood and adolescence (F94) | 996 (0.1) | 0 (0.0) | 0 (0.0) | 0 (0.0) | 996 (0.0) | 1 194 (0.2) | 0 (0.0) | 0 (0.0) | 0 (0.0) | 1 194 (0.0) |
| Tic disorders (F95) | 475 (0.1) | 57 (0.0) | 0 (0.0) | 0 (0.0) | 532 (0.0) | 1 771 (0.2) | 131 (0.0) | 12 (0.0) | 0 (0.0) | 1 914 (0.1) |
| Other behavioral and emotional disorders with onset usually occurring in childhood and adolescence (F98) | 3 338 (0.5) | 163 (0.0) | 10 (0.0) | 15 (0.0) | 3 526 (0.1) | 5 416 (0.7) | 153 (0.0) | 13 (0.0) | 0 (0.0) | 5 582 (0.2) |
| Unspecified mental disorder (F99) | 536 (0.1) | 863 (0.1) | 378 (0.0) | 74 (0.0) | 1 851 (0.1) | 373 (0.0) | 629 (0.1) | 282 (0.0) | 27 (0.0) | 1 311 (0.0) |
| ^1^Service utilization is the number of individuals with any medical contacts with a diagnosis of a mental disorders during the year 2019, divided by the number of individuals in the study population on December 31, 2019. | | | | | | | | | | |
